# Climate extremes increase dengue risk along elevation and socio-economic gradients in Colombia

**DOI:** 10.1101/2024.04.02.24304484

**Authors:** Pallavi Kache, Daniel Ruiz-Carrascal, Rachel Lowe, Anna M. Stewart-Ibarra, Karen C. Seto, Maria Diuk-Wasser, Mauricio Santos-Vega

**Affiliations:** Department of Ecology, Evolution, and Environmental Biology, Columbia University, New York, USA; International Research Institute for Climate and Society, Columbia University, New York, USA; Barcelona Supercomputing Center (BSC), Barcelona, Spain; Catalan Institution for Research and Advanced Studies (ICREA), Barcelona, Spain; Centre on Climate Change & Planetary Health and Centre for Mathematical Modelling of Infectious Diseases, London School of Hygiene & Tropical Medicine, London, UK; InterAmerican Institute for Global Change Research (IAI). Montevideo, Uruguay; Yale School of the Environment, Yale University. New Haven, USA; Grupo Biología Matemática y Computacional, Departamento de Ingeniería Biomédica, Universidad de los Andes, Bogotá D.C., Colombia; Departamento de Ciencias Biológicas, Universidad de los Andes, Bogotá D.C., Colombia

**Author notes:** **CORRESPONDING AUTHOR**: Pallavi A. Kache 1200 Amsterdam Ave. New York, NY 10027. Equal contributions as senior author (Maria Diuk-Wasser, Mauricio Santos-Vega).

## Abstract

Globally, urban settlements face increases in the frequency, magnitude, and duration of extreme climate events and shifts in their timing and spatial extent. Variation in temperature and rainfall conditions affect the temporal onset of dengue transmission. However, there is a need to understand how climate-related patterns and disease transmission mechanisms vary by location, particularly for topographically complex landscapes. In this investigation, we used dengue cases from 1,120 municipalities and five regions across Colombia during 2008–2019, and analyzed associations with extreme climate covariates generated from fine-scale, daily-level meteorological data, accounting for varying landscape and socio-economic properties. Using Bayesian spatio-temporal hierarchical models, we determined that high-intensity warm spells (with positive temperature anomalies of 8–12°C above mean monthly conditions) resulted in an earlier onset of dengue transmission risk in high-elevation settlements compared to low-elevation settlements. Furthermore, the risk of dengue transmission after extremely dry conditions was greater and extended for a longer duration in highly urbanized municipalities compared to those with a low urban population. Our findings highlight that meteorological hazards affect disease transmission in urban settlements differently based on elevation and socio-economic conditions. Additionally, our analysis adds to increasing evidence of the vulnerability of mountainous urban communities to extreme weather and vector-borne diseases. Overall, we emphasize the need for monitoring and forecasting the occurrence and intensity of meteorological hazards and associations with emerging climate-sensitive disease threats.

## Main

Globally, urban settlements are facing increases in the frequency, magnitude, and duration of extreme climate events, as well as shifts in their timing and spatial extent, associated with climate change^1,2^. Heavy rainfall, droughts, and warm spells pose environmental hazards and exacerbate the existing challenges that urban populations face, including housing security, water supply, and socio-economic inequalities^3,4^. These inter-linked climate and urban issues directly and indirectly impact human health, including infectious disease transmission. Vector-borne diseases are particularly sensitive to climate conditions^5^. Temperature drives the biology of vectors and pathogens, and rainfall (as well as human water storage and management practices) regulates the availability of standing water where mosquitoes lay their eggs, determining the size and distribution of mosquito populations. Nearly half of the global population is at risk for dengue virus (DENV 1-4) infection—living in urban centers with the implicated *Aedes* spp. mosquitoes present and under environmental conditions suitable for virus transmission^6,7^. *Aedes* spp. mosquitoes feed on humans as a blood meal source and lay their eggs in water-holding containers, including discarded waste and water storage receptacles found across human-dominated landscapes^8^. Therefore, climate interacts with the socio-economic conditions of an urban settlement (e.g., human population density, availability of waste and water management) to determine dengue risk^9^.

Temperature and rainfall act on multiple co-occurring and interacting processes that drive DENV transmission. As a result, climate affects dengue risk in complex and non-linear ways that vary over space and time, often with time lags that range from several weeks to months^10,11^. Ambient air temperature determines the speed of *Aedes* spp. life history and behavioral processes (e.g., development rates, survival rates, biting rates, etc.) as well as DENV replication rates within the mosquito. DENV transmission increases with rising temperatures, peaking at mean temperatures of 26–29°C, and then diminishes above this thermal optimum^12^. Rainfall leads to water accumulation in containers and infrastructure found throughout urban environments, creating aquatic habitat for juvenile mosquito development. Human water storage practices further complicates these dynamics, as people living in water-scarce regions (or settlements with unreliable water access) store water in containers around the home and community, that serve as egg-laying sites for female mosquitoes.

Over two decades of climate and dengue research has established the associations between inter-annual climate phenomena (e.g., El Niño-Southern Oscillation, ENSO) and monthly weather conditions (e.g., mean monthly temperature) on dengue risk^5,13–17^. The association between extreme climate events and dengue, however, remains comparatively understudied. Evidence suggests that outbreak risk increases one to three months after heatwave events^18,19^. With respect to rainfall, heavy rains may flush out juvenile populations from water-holding containers, decreasing initial risk^20^. During dry spells, the need for communities to store water (for daily household uses such as cleaning and cooking) contributes to a build-up of aquatic habitat and mosquito populations over time, with studies showing spikes in dengue incidence three to five months after the onset of dry conditions^5^. However, the nuanced effects of extreme climate events, with different intensities and durations, on dengue risk requires further investigation.

Urban settlements are not uniformly vulnerable to the impacts of climate extremes on disease risk^21^. Certain regions, for example highland areas, are particularly sensitive to changes in temperature^22^. These cooler environments at the limits of the historical range of *Aedes*-borne diseases, are increasingly vulnerable to emerging disease outbreaks associated with warming temperatures^23^. In Puerto Rico, for example, temperature variability had the greatest impact on dengue incidence in the island’s mountainous zones^24^. In Nepal, climate change has intensified the magnitude of dengue outbreaks in mountain regions^25,26^.

The present-day socio-economic characteristics of urban settlements also determine how climate extremes impact dengue transmission risk. For example, in resource-limited settings, such as large informal urban settlements, heavy rainfall events coupled with garbage accumulation and poor housing conditions may result in the creation of high densities of aquatic larval habitat within a short span of time. Whereas, settlements that are formally- constructed with high levels of impervious cover, may see higher rates of flash flooding and washout of larval habitat. These hypothesized interactions between climate conditions and urban features are generated from a limited number of studies. To inform dengue prevention in the midst of a changing climate, there is a need to understand how climate extremes drives dengue risk for urban settlements with different geographic features and levels of socio-economic vulnerability.

Here, we highlight the country of Colombia as a case study to examine the nexus of extreme climate, urban socio-economic conditions, and dengue. Colombia is a highly urban country and labeled as “megadiverse,” containing a high degree of spatial heterogeneity in climate regimes and ecosystems^27^. The country, therefore, offers immense opportunity to examine how climate extremes interact with the geography and socio-economic conditions of urban settlements to drive dengue risk. We coupled spatio-temporal Bayesian hierarchical models with distributed lagged non-linear models (DLNMs). This approach simultaneously quantifies the spatially-varying, non-linear, and delayed dependencies between dengue cases and extreme climate conditions^28^. We use fine-scale, daily-level data to derive extreme climate metrics, emphasizing how short-term fluctuations in meteorological conditions have long-lasting impacts on dengue risk. Given Colombia’s high degree of heterogeneity, results from this study can be used to inform the development of region-specific dengue early warning systems and outbreak preparedness activities for urban settlements across South America and the world.

### Dengue, Extreme Weather, and Urban Characteristics

Our study included 999,523 cases of dengue across 1,120 municipalities in Colombia from January 2008 to December 2019. National outbreaks occurred in 2010, 2013, 2016, and 2019 (Fig. 1a). Dengue incidence was highest for departments in the Andean, Orinoco, and Amazon regions (Fig. 1b). Dengue seasonality varied along the country’s coastline, with higher incidence during the first half of the year on the Pacific Coast, and during the second half on the Caribbean Coast. For interior territories, the Orinoco and Amazon, dengue incidence was higher in the first to middle months of the year. In the Andean region, departments with municipalities at lower elevations (e.g., Tolima, Huila, Norte de Santander) experienced high incidence year-round and consistently during the study period. In contrast, those at higher elevations (e.g., Cundinamarca, Boyacá) exhibited high rates only during outbreak years (Fig 1b, Fig. S2). Through field sampling, *Aedes* spp. mosquitoes have been detected at elevations as high as 2,300 meters, with mosquitoes testing positive for dengue virus up to 1,900 meters^29,30^. Here, we observed cases in 44 municipalities with elevations above 2,300 meters, with cases detected as high as 3,165 meters. Cases at the highest end of this elevation range are hypothesized to be travel-associated, with infections acquired at lower-elevation municipalities.

**Fig 1.**
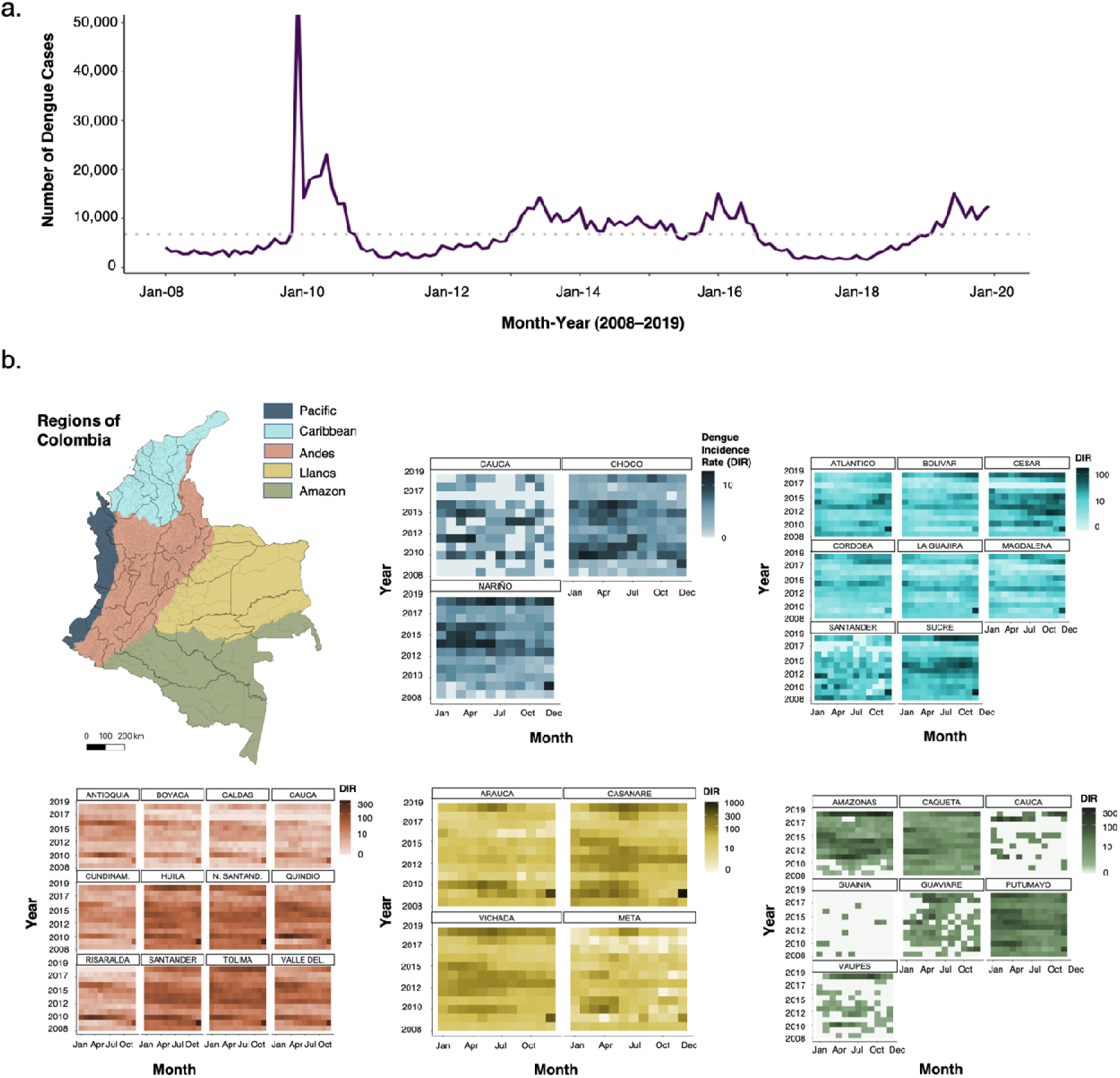
Dengue cases in Colombia, 2008–2016. **a**. Timeseries of dengue case counts for Colombia from January 2008– December 2019. The horizontal line represents the average number of dengue cases per month for the study period (N=6704). **b.** Heatmaps of dengue incidence rates (DIR) for each department, grouped and colored by region. The scales vary to highlight inter-annual and seasonal variations in DIR within and between regions. Additionally, some departments span multiple regions (e.g., Cauca spans both the Andean and Pacific regions); data reflect the sum of dengue cases reported in the municipalities within the indicated region.

Measured climate extremes were frequent during our study period, consistent with evidence that Colombia is highly vulnerable to extreme climate, particularly during ENSO events^31,32^. Approximately 58% of municipalities (N=648/1,120) experienced at least one month where monthly temperature anomalies, precipitation anomalies, or both that exceeded the upper tercile their long-term average (1981–2019) (Fig 2a). Additionally, temperature anomalies exhibited elevation dependency for the 594 municipalities between 500–2500 meters. For example, municipalities between 1000–1500 meters experienced average temperature anomalies of 1.04°C, those at 1500– 2000 meters, 1.10°C, and those between 2000–2500 meters, 1.13°C (Fig 3).

**Fig 2.**
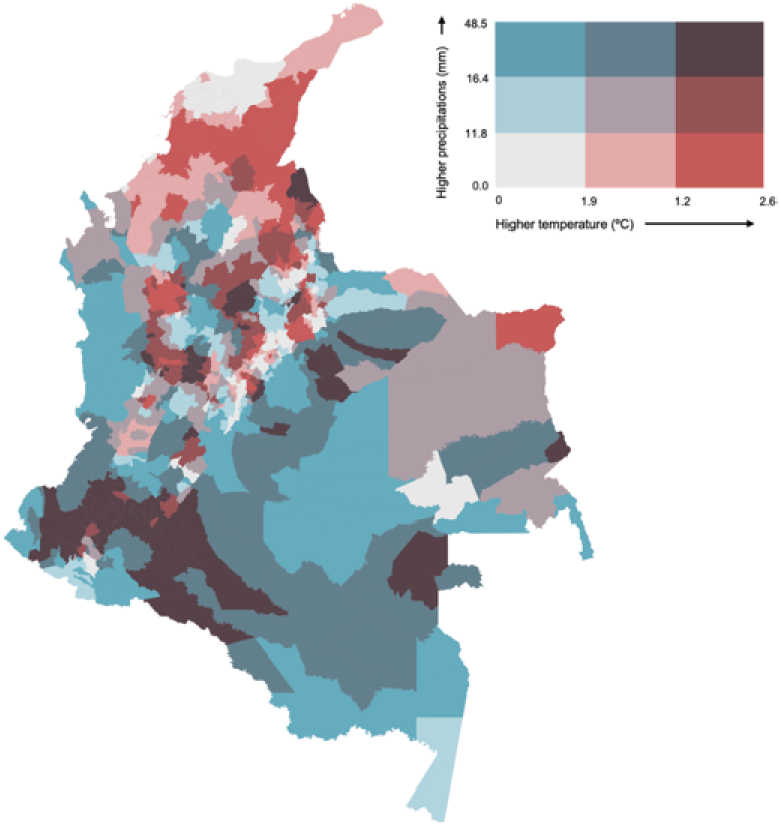
Bivariate plots of extreme temperature and extreme rainfall conditions. a. Bivariate plot showing terciles of mean monthly temperature anomalies and excess rainfall anomalies, compared to their long-term average (1981–2019), averaged across the study period (2008–2019).

**Fig 3.**
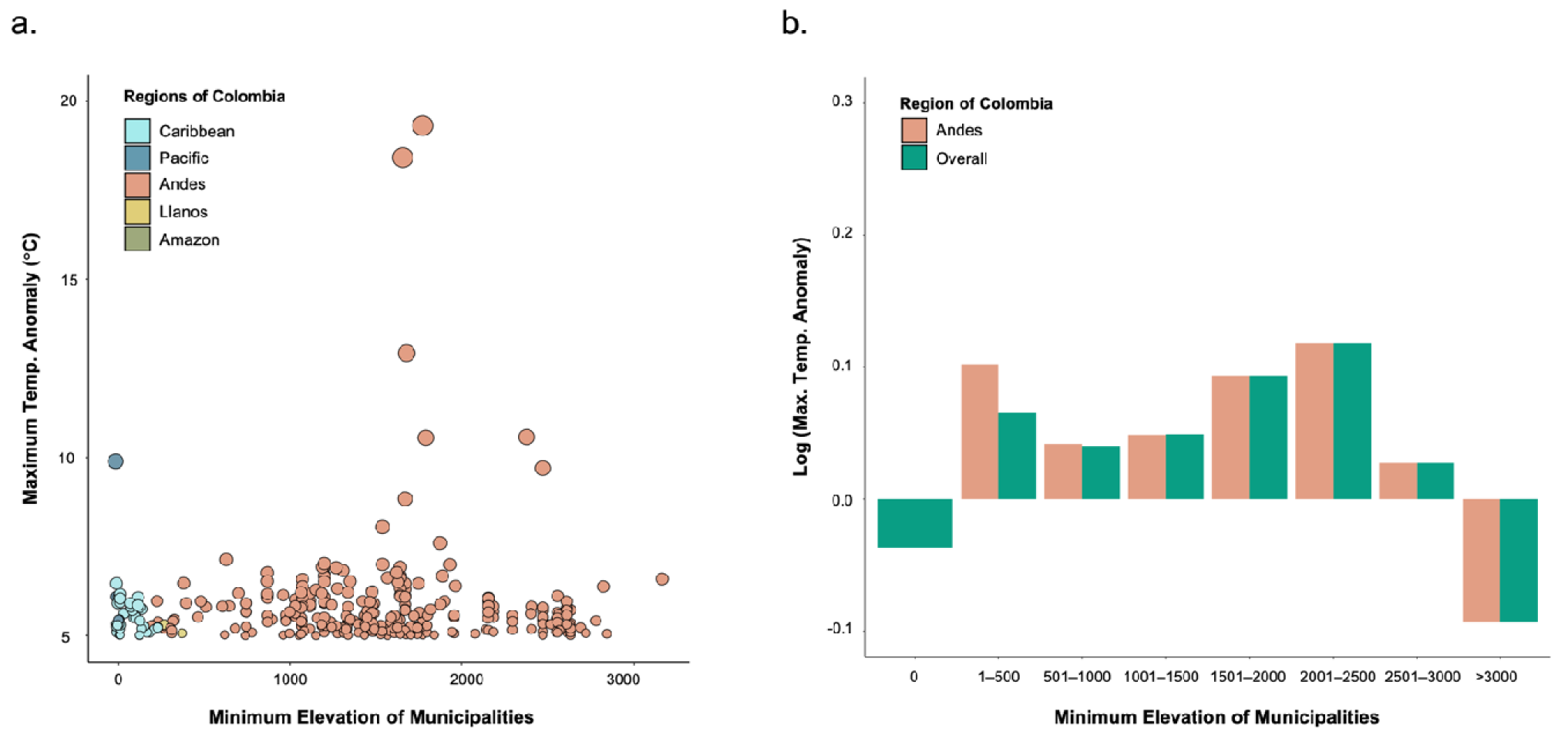
Association between maximum temperature anomalies and municipality elevation. **a**. Scatterplot of maximum temperature anomalies above 5°C and the minimum elevation of municipalities, colored by region. **b.** Bar chart of the log of maximum temperature anomalies and elevation, binned in increments of 500 meters. Bars are shown for Colombia overall and for the Andes region.

Half of Colombia’s municipalities have 40% or more of their population residing in urban settlements. Municipalities are distributed along a wide elevation range (0–3,165 meters), with weak associations between elevation and the percent urban (r=-0.23), indicating that the proportion of the population living in urban settlements is not elevation-dependent. There are strong positive correlations between the percent urban and access to water systems (r=0.77); however only moderate negative associations between the percent urban and the proportion “living with adequate housing conditions” (r=-0.4), as measured by the 2018 census^33^.

### Comparing Models of Monthly Climate Extremes to Models of Mean Conditions

As a first step, we established a baseline model to predict monthly relative risk (RR) of dengue, with year-specific municipality-level spatial random effects, monthly department-level temporal random effects, and a regional fixed effect (Table S2). We then added to this baseline by comparing dengue-climate models of monthly climate extremes to models of mean conditions—with lagged between 0 and 6 months for each climate variable. We explored extreme temperature variables including the number of warm days per month (WarmDays; i.e., days exceeding the 90th percentile of long-term mean conditions, T_90-mean_); the maximum duration of a warm spell (MaxDuration_WarmSpell_; i.e., maximum duration of consecutive warm days); and the maximum intensity of a warm spell (MaxIntensity_WarmSpell_; i.e., maximum positive temperature anomaly (°C) of a warm spell over long-term mean conditions). We also explored extreme precipitation variables including monthly positive precipitation anomaly (Anomaly_Precip_, i.e., exceedance of monthly rainfall over the monthly long-term mean); the number dry and wet days (DryDays, WetDays; i.e. days with less than or greater than 1 mm of precipitation, respectively); and the maximum dry and wet spell durations (MaxDuration_DrySpell_, MaxDuration_WetSpell_). Full definitions of extreme climate variables are provided in Table 1.

**Table 1.**
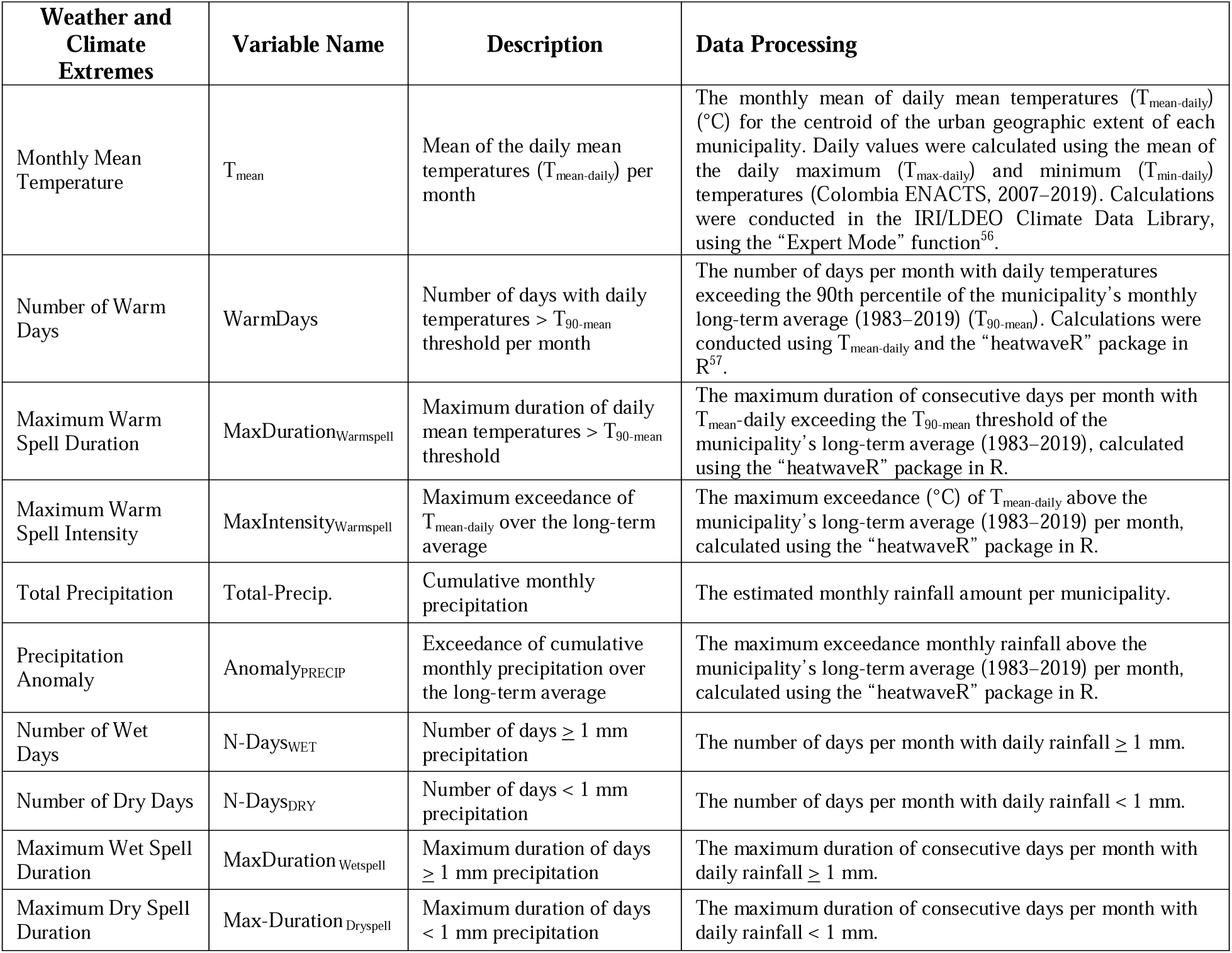
Monthly weather and climate extremes variables calculated from daily-level data.

For temperature, we found that the T_mean_ model had a lower DIC than three extreme temperature models (WarmDays, MaxDuration_WarmSpell_, MaxIntensity_WarmSpell_). However, the mean absolute error (MAE) of the MaxIntensity_WarmSpell_ model was lower than the MAE of the T_mean_ model for 26% of municipalities across the country, indicating better prediction in these areas (Table S5, Fig. S6–S7). We observed this improved model prediction for 25% of municipalities in the Caribbean, 28% in the Andean, and 34% in the Pacific region (Fig. S6). Within the Andean region, improved model prediction was indicated for 29% of intermediate- and 25% of high-elevation municipalities, compared to 11% of low-elevation municipalities. Therefore, we examined the additive and interactive effects of MaxIntensity_WarmSpell_ and municipality elevation on lagged dengue risk. The interaction model (MaxIntensity_WarmSpell_ *Elevation) had a substantially improved model fit over the T_mean_ model (ΔDIC = 10949).

For dry conditions, the DryDays model had a better fit than the Total-Precip model (ΔDIC = 301). We observed an improvement using DryDays over Total-Precip for 48% of municipalities across the country, with the greatest added value seen for municipalities in the Andean, Amazon, and Orinoco regions (with added value for 51%, 66%, and 80% of municipalities in the regions, respectively). Next, we explored the interaction between DryDays and the probability that individuals may need to store water within the household. We hypothesized that variables including the proportion of residents with water system access or inadequate household conditions, would more closely reflect the need for water storage and yield the best model fit; however, this was not reflected in the data. Instead, the model with percent of the population residing in urban settlements (i.e., Percent Urban) yielded a substantially better fit over the proportion of residents with water system access model (ΔDIC = 5084).

Finally, we found that the number of rainy days per month (WetDays) had a better fit than Total-Precip (ΔDIC = 114) and other measures of extreme precipitation, including monthly precipitation anomalies (ΔDIC = 691). We observed an improvement using WetDays over Total-Precip for 55% of municipalities across the country, with the greatest added value seen for municipalities in the Andes, Amazon, and Orinoco regions (with added value for 55%, 72%, and 77% of municipalities in the regions, respectively). We hypothesized that variables including human population density or the percentage of the municipality population residing in low-income housing would more closely reflect the potential for water container accumulation in the urban landscape; however, the interaction model including WetDays *Percent Urban best fit the data. Given that the best-fit models for rainfall conditions included the interaction between wet or dry days with the percent urban, we constructed one final model, incorporating the cumulative effects of extreme temperature-elevation interactions and rainfall conditions-urban interactions (Table 3).

### Warm spells and Dengue Risk along an Elevation Gradient

Warm spells had a sustained impact on the RR of dengue for up to six months (Fig. S12). From the best-fitting model (MaxIntensity_Warmspell_), we found that warm spells (> 2 days) with positive temperature anomalies (i.e., intensities) of 12–13°C above the municipality’s long-term mean temperature contributed to a RR peak 2–4 months after the extreme temperature event occurrence (RR > 4.0) (Fig. S12). When observing MaxIntensity_Warmspell_ along an elevation gradient, we made two observations regarding the temporality of the lagged response and the maximum RR (Fig 4). We found that at low elevations, the maximum RR occurred 2–3 months after an anomalous temperature event. However, for municipalities above 1,000 meters, RR was highest in the first two months after an extreme temperature event (Fig. 4, Table 3). Furthermore, at higher elevations, moderate-intensity warm spells (2–6°C) resulted in sustained RR for the full six-month observation period, whereas high-intensity temperature anomalies (8– 12°C) contributed to increases in RR for a slightly shorter period (for up to five months, Fig S14). As anticipated, the maximum RR was higher for low-elevation municipalities compared to high-elevation municipalities, given that ambient air temperatures decrease at higher elevations. For low-elevation municipalities, the maximum RR exceeded 4.0; for municipalities above 1,500 meters, maximum RR was below 3.0, and at the highest elevations (∼2,500 meters) the maximum RR stayed below 2.0 (Fig. 4, Fig. S11).

**Fig 4.**
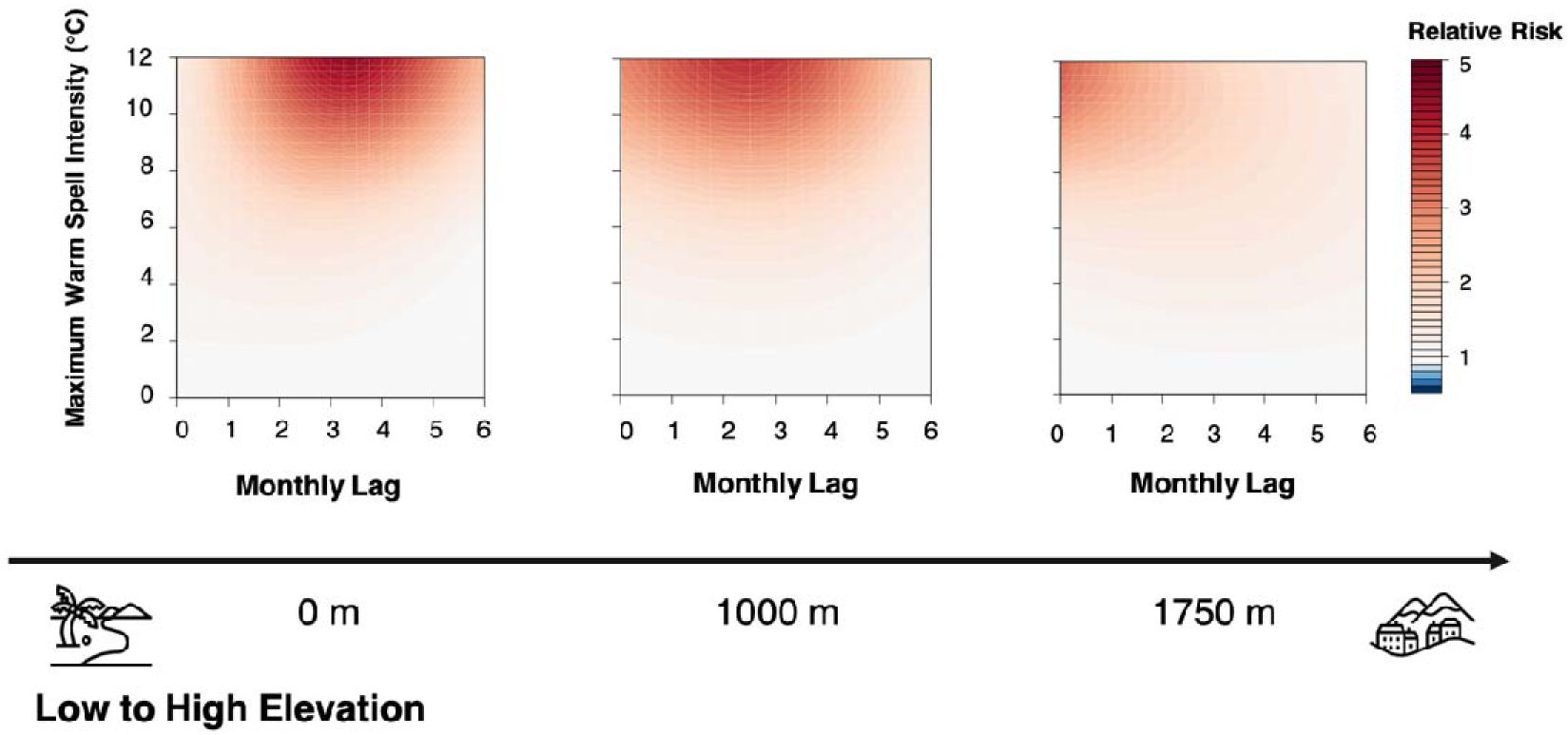
Warm spell intensities and dengue risk, along an elevation gradient. Contour plots show the exposure-lag-response association between the maximum warm spell intensity in a month and dengue using the temperature and precipitation interaction model. Darker red indicates an increase in the relative risk of dengue compared to monthly normal temperatures (calculated from monthly averages, 1981–2019). Plots are restricted

**Fig 5.**
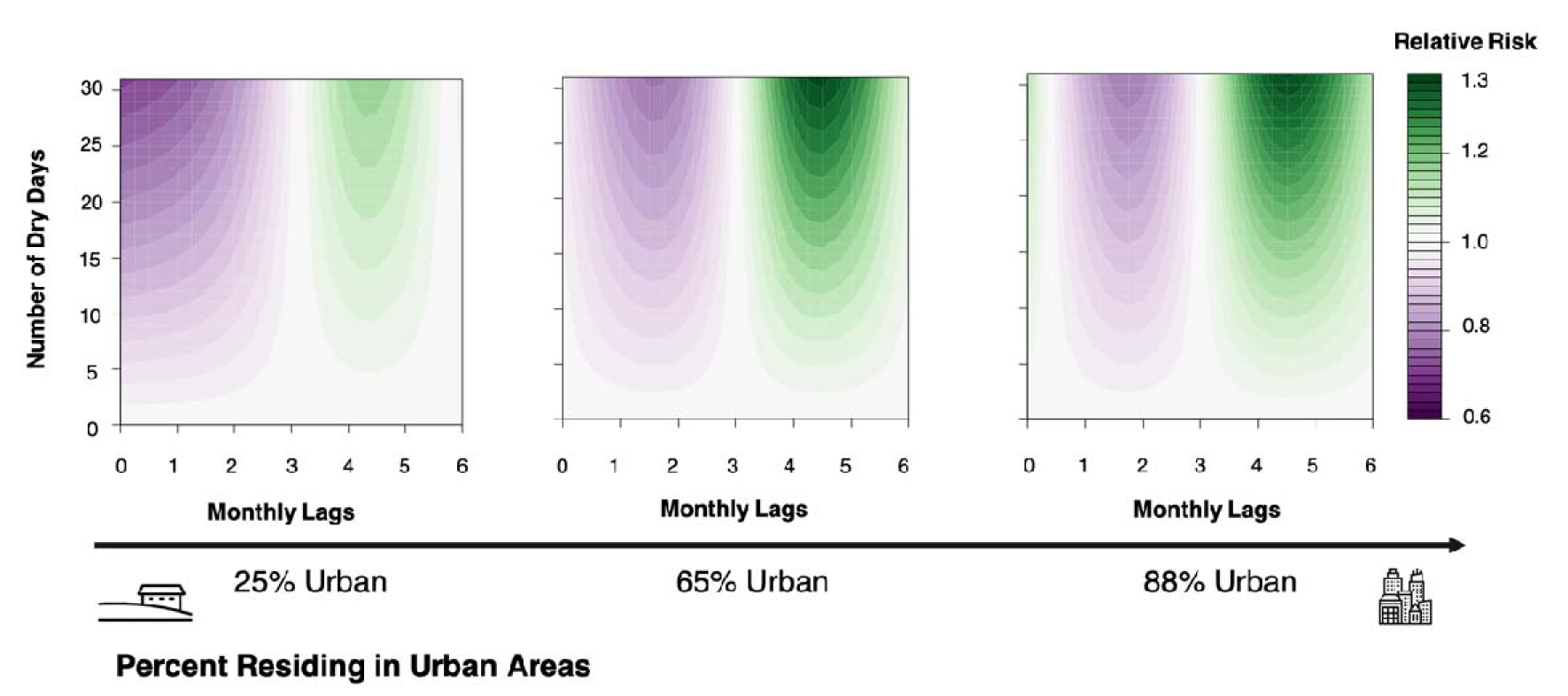
Occurrence of dry days and dengue risk, along a percent urban gradient. Contour plots show the exposure-lag-response association between extreme temperature anomalies and dengue using the dry conditions-percent urban model. The darker shade of teal indicates an increase in the relative risk of dengue compared to zero dry days.

Other extreme temperature indicators provided information on the exposure-lag-response relationship with dengue incidence and served as points of comparison for previous dengue DLNM studies. Models of warm spell duration (MaxDuration_WarmSpell_) indicated that a warm spell of ≥ 2 days is associated with increases in RR for up to six months (Fig. S16a). When this warm spell lasts 15 days, the highest risk (RR>1.25) is observed within the first three months before decreasing over time; and when a warm spell lasts nearly one month (>25 days), the highest risk (RR>1.4) is observed within a matter of weeks before decreasing over time.

### Extreme Rainfall and Dengue Risk along a Socio-economic Gradient (280)

When examining the effects of dry days on dengue risk, across levels of urbanization, there is an initial decrease in risk within the first 0–2 months, followed by an increase in risk at 3–6 months. Increases in risk (i.e., RR>1) were triggered by as few as 3–5 dry days per month. The highest levels of risk were observed when almost the entire month was made up of dry days (≥ 26 dry days), with a peak RR occurring at a 4–5-month lag.

When examining this rainfall-dengue relationship along a gradient of less to more urbanized (based on percent urban), RR was higher for highly urbanized municipalities (≥ 60% of the population living in urban settlements) compared to less urbanized municipalities (≤25% urban), and this risk extended for a longer duration of time. For instance, less urbanized municipalities experienced maximum dengue risk (RR=1.1) with a 3–5-month lag, whereas highly urban municipalities (60% urban) experienced a RR of 1.4, with risk extending up to six months. Similarly, the initial protective effect of dry conditions are greater for rural municipalities than urban municipalities, and happen more immediately. Specifically, for rural municipalities, the lowest RR is 0.6 and occurs within the first two months of a municipality experiencing dry conditions. For highly urban municipalities, the lowest RR is 0.82 and occurs with a lag of 1–3 months after a municipality experiences dry conditions.

## Discussion

In this study, we examined the lagged and non-linear effects of climate extremes on dengue risk in Colombia, a highly urban country with a topographically-complex landscape. We found that moderate-intensity warm spells extended the duration of increased dengue risk up to six months. High-intensity warm spells posed a more immediate (i.e., short-term) risk for high-elevation municipalities than low-elevation municipalities. Additionally, our results support previous indications that dry conditions contribute to increased dengue risk at a 3– 5-month time lag. The magnitude of risk is higher for urban municipalities than rural municipalities^28^. Our investigation uniquely tested monthly-level variables of extreme climate derived from daily-level weather data (blended from satellite imagery and weather station data), revealing promising, and thus far under-utilized, indicators for future climate and vector-borne disease investigations.

The greatest temperature anomalies were observed in the Andean region, prompting us to investigate the interactive effects of extreme temperature and elevation on dengue risk. While previous studies have examined the effects of extreme temperature on dengue risk, the effect modification of this risk by elevation has not previously been quantified^18,19,34^. Here, we found that high-elevation settlements (>1,750 meters) had an immediate increase in dengue relative risk after a high-intensity warm spell (8–12°C above normal monthly conditions) and an extended duration of the highest-risk period. However, low-elevation settlements below (<1,000 meters), had the highest dengue relative risk 2–3 months after a high-intensity warm spell.

We attribute these differences to ambient temperature conditions of the municipality, and in particular, where the municipality is situated along the thermal response curve for dengue virus transmission probability. Laboratory and modeling studies have indicated that for the dengue virus, optimal transmission conditions occur between 26–29°C; above this, the transmission probability decreases^12^. For our study period, high-elevation Colombian municipalities had median average annual temperatures of 15°C (IQR: 13°C, 18°C), while low-elevation municipalities had median average annual temperature of 27°C (IQR: 25°C, 28°C). Therefore, for high-elevation municipalities with established *Aedes* spp. populations, vector and virus traits are optimized during warm spell intensities of 8–12°C, whereas for low-elevation municipalities, these biological processes would exceed their thermal optimum.

Our findings highlight the value of tailoring climate information into meaningful indicators of vector biological and ecological processes, as opposed to relying on mean conditions alone. An entomological investigation by Carrington et al. (2014) showed that temperature fluctuations in aquatic larval habitats influence immature mosquito development, while changes in ambient air temperature affected female egg production (i.e., fecundity)^35^. Specifically, large diurnal temperature variation (18.6°C) at low-temperature water conditions (16°C) increased larval survival and decreased development time compared to constant temperature conditions^36,37^. However, small fluctuations (7.6°C) at intermediate and high mean temperatures (26°C and 35°C, respectively) slowed the development of immature mosquitoes^35^ Furthermore, they demonstrated that small variations in daily ambient temperature at high mean temperatures terminated female egg production^35^. While daily temperature variation of aquatic habitat is not directly associated with the warm spell intensities that we used in our study, these experimental findings provide supportive evidence that juvenile development may accelerate under extreme temperature conditions at high elevations, resulting in increased dengue risk within a short time window. In contrast, juvenile development may slow down and female mosquito fecundity may decrease under these same extreme temperatures at low elevations, contributing to delayed increases in dengue incidence.

For low-elevation regions with high baseline temperatures, a study in Vietnam also described a delayed spike in dengue risk after a heatwave (defined as more than seven days of temperatures > 95^th^ percentile, 24.3°C)^19^. While in Singapore, researchers found initial decreases in dengue relative risk (RR<1) in the weeks immediately following heatwaves (three or more days of temperatures > 90^th^ percentile, 33.2°C)^32^. Alternatively, in attempts to explain the demographic mechanisms behind delayed mosquito population outbreaks at anomalously high temperatures, Chaves et al. 2014 implemented a two-stage demographic model of *Ae. aegypti* larvae and adults^38^. They observed 10-week delays as high-temperature environments initially triggered decreases in larval survival, but later initiated compensatory increases in fecundity (which were assumed within the model, based on population fitness trade-offs) that lasted the span of the heatwave, as well as increases in larval survival and responsive decreases in fecundity when temperature conditions returned to normal. While there are indications that extreme temperature anomalies at high mean temperatures delay the onset of increased risk, the biological and statistical links between extreme temperature conditions and mosquito ecology are poorly understood. Further laboratory, field, and statistical investigations are needed^5,39^.

Our findings indicate a different dengue risk profile for high versus low-elevation municipalities under extreme temperature conditions. First, high-elevation municipalities experiencing anomalous temperatures may not have the same early warning lead time for a dengue outbreak as low-elevation municipalities. Additionally, cities at high elevations likely have a larger proportion of susceptible individuals in their population (due to a lack of endemic circulation of DENV), which may contribute to an earlier onset of pathogen transmission^40,41^. Furthermore, high-elevation municipalities that do not commonly experience dengue circulation may not have the necessary public health infrastructure to promptly respond to emerging outbreaks, further increasing population-level vulnerability to the disease. This is particularly true for a country such as Colombia, where many people reside in small- to mid-sized urban settlements at high elevations. While warm spells in low-elevation regions carry public health risks due to heat stress and associated non-communicable disease risks, proactive interventions including dengue advisories should also be initiated for high elevation urban settlements to prevent rapid-onset dengue outbreaks^42,43^.

With respect to precipitation extremes, urban municipalities experienced a maximum RR of 1.4 between three to six months after a high occurrence of dry days per month. Rural municipalities, however, experienced a maximum RR of 1.2 between three to five months after a high occurrence of dry days per month. The increased risk in urban environments may be due to higher mosquito populations densities in urban environments compared to rural ones, due to the availability of larval habitat and human blood-meal hosts. Additionally, highly urban municipalities may have a strained capacity to ensure water security for a large population, necessitating the collection and storage of water in artificial containers for a longer period of time compared to rural areas^28^. Here, the temporal lag in dengue risk following dry conditions is similar to previous national-level investigations conducted by Lowe et al. for Barbados and Brazil^10,28^. The extended duration of risk for Colombian urban settlements following dry conditions may be due to water management and governance factors, socio-economic conditions, or behavioral practices that leave urban Colombian residents vulnerable to dengue risk for a longer time period.

As research on extreme weather and dengue risk continues, we find that a systematic evaluation of precipitation indicators is needed, particularly in assessing risk at multiple spatial and temporal scales or for different decision-making criteria. Notably, we found that a simple measure of month-to-month count of days without rainfall resulted in a similar temporal lag in increased dengue incidence compared to drought indices used in other studies. This variable may serve as a more tractable weather indicator for policy-makers and a user community—easy to understand and calculate using readily-available data^44^. Specifically, we calculated a crude measure of monthly dry conditions (the frequency of dry days per month), whereas previous national-level investigations conducted by Lowe et al., leveraged more sophisticated drought indices such as the Palmer Drought Severity Index (PDSI) or the Standardized Precipitation Index (SPI)^10,28^. These indices incorporate normal precipitation conditions using historical records, and in the case of SPI, consider dry conditions across multiple time horizons, ranging from one month (SPI-1) to 24 months (SPI-24), which capture prolonged dry spells or drought.

Our study supports the development of early warning systems by quantifying a specific time window following extreme weather conditions, during which outbreak preparedness activities can be initiated. These measures may include enhancing mosquito control, preparing health facility capacity, and communicating with local communities regarding the increased risk of dengue transmission. We emphasize that weather hazards can affect urban settlements differently based on elevation and socio-economic conditions. Additionally, our analysis adds to increasing evidence of the vulnerability of mountainous urban communities to extreme weather and vector-borne diseases, emphasizing the need for monitoring and forecasting the occurrence and intensity of meteorological hazards and their associated health impacts.

## Methods

### Data Sources

We obtained dengue case data from 2008–2019 using the national surveillance system, SIVIGILA (*Sistema Nacional de Vigilancia en Salud Pública*), operated by the National Institute of Health (*Instituto Nacional de Salud*). To calculate dengue incidence rates per municipality, we divided monthly case counts by annual population estimates using census data collected in 2005 and 2018 (for years 2008–2017 and 2018–2019, respectively)^45,46^.

Temperature data were obtained from the Colombia’s Enhancing National Climate Services (ENACTS) initiative of the International Research Institute for Climate and Society (IRI)^47^. The dataset provided daily temperatures from 1980–2019 at a 0.1° spatial resolution. Data were blended (i.e., spatially and temporally interpolated and smoothed) using NASA’s MERRA-2 data and ground-truth records gathered at weather stations operated by the Colombian Institute of Hydrology, Meteorology and Environmental Studies (*Instituto de Hidrología, Meteorología y Estudios Ambientales*, IDEAM)^48^. For each municipality, we used the mean of daily minimum and maximum temperatures to calculate a daily mean temperature (T_mean-daily_). We then used T_mean-daily_ to derive mean monthly temperatures (T_mean_) and monthly extreme temperature variables, including the total number of warm days per month (days with temperatures exceeding the 90^th^ percentile of the municipality’s long-term seasonal climatology, 1980–2019, T_90-mean_), the maximum duration a warm spell for a month (the maximum number of consecutive days with temperatures exceeding the T_90-mean_), and the maximum warm spell intensity per month (positive temperature anomaly of warm spell compared to the long-term climatology, 1980–2019).

We also obtained daily precipitation data from the Climate Hazards Group Infrared Precipitation with Station data (CHIRPS)^49^. CHIRPS provides precipitation estimates from 1981–2019 at a 0.05° spatial resolution^49^. With these data we calculated monthly total precipitation amounts, the monthly precipitation anomaly, the longest wet spell duration (duration of consecutive days with > 1mm daily rainfall), the longest dry spell duration (duration of consecutive days with < 1mm daily rainfall), and the number of dry days (number of days with < 1mm daily rainfall) (Table 1).

We examined how extreme climate variables interact with each urban settlement’s geographic and socio- economic characteristics to drive dengue incidence. Each municipality’s region and Lang climate zone designations were obtained using IDEAM shapefiles^50^. Additionally, we calculated the minimum elevation of the urban center for each municipality with data from the Shuttle Radar Topography Mission (SRTM v3 product) at a 30-meter resolution (S1 Text)^51^. To characterize the socio-economic characteristics of each municipality, we used cross-sectional data taken from the 2018 national census, considering the percentage of residents living in urban areas (i.e., the percent urban), population density, the percentage of residents with water system access, and the percentage of homes with adequate housing conditions^45^. We ran a principal components analysis (PCA) to understand the total variability among elevation and urban socio-economic characteristics (Fig. S5).

### Modeling Approach

We constructed spatio-temporal Bayesian hierarchical models of monthly dengue case counts for 1,120 Colombian municipalities in 31 departments from January 2008 to December 2019. We used negative binomial regressions to account for overdispersion in the distribution of case counts (S1 Text, Fig. S3). We incorporated a distributed lagged nonlinear model (DLNM) to examine non-linear and delayed associations between climate extremes and dengue incidence from 0 to 6 months, as indicated from previous investigations^10^. Model parameters were estimated in a Bayesian framework using integrated nested Laplace approximations in R version 4.2.1 (S1 Text) ^52^.

We constructed baseline models with department-level monthly autocorrelated random effects to account for seasonal variation. Additionally, baseline models included municipality-level yearly spatial random effects to account for inter-annual variation in shared attributes between neighboring municipalities (e.g., human mobility) or other factors not included in our analysis (e.g., dengue serotypes in circulation, vector control activities). We then tested whether including five regions and six Lang climate classifications as fixed effects improved the model fit. We incorporated DLNMs for the weather and climate extremes covariates to understand possible non-linear and delayed associations between dengue incidence, extreme temperature, and rainfall from 0 to 6 months.

We tested how extreme temperature and rainfall indices interact with elevation and socio-economic characteristics to drive dengue incidence. For temperature, given the evidence for elevation-dependent warming in the Andes Mountains, we tested the linear interaction between extreme temperature variables and the minimum elevation of the urban extent for each municipality^53^. We centered the elevation variable at 0 meters, 1,000 meters, and 1,750 meters to partition low-, intermediate-, and high-elevation municipalities.

For dry conditions, based on previously established associations between drought and household water storage on dengue risk, we examined whether socio-economic variables might serve as a proxy for the population-level need for water storage^28^. We tested the interactions between dry conditions and socio-economic variables using linear interaction terms by centering the continuous variables at low, intermediate, and high values of each variable (Table 2). We first tested the percent of the population living in an urban area. We then examined the municipality’s water infrastructure level, measured by the percentage of residents accessing water systems, and finally, tested whether household-level properties may reveal the need for water storage, using a summary index calculated from the 2018 Colombian national census measuring a lack of “household deficiencies” (which included questions regarding access to public services and the structural stability of the dwelling)^33^. For excess rainfall conditions, we aimed to capture the interaction between heavy rainfall and the availability of water-holding containers in the urban landscape. We used linear interaction terms for socio-economic characteristics, including the percent urban, population density, and the percent of the population living in low socio-economic status neighborhoods. Similarly to dry conditions, we centered these continuous variables at low, intermediate, and high values (Table 2).

**Table 2.** Centered socio-economic variables for climate extremes-urbanicity interaction models^‡^.

| Socio-economic Condition | Centered: Low |  | Centered: Intermediate |  | Centered: High |  |
| --- | --- | --- | --- | --- | --- | --- |
|  | Percentile | Value | Percentile | Value | Percentile | Value |
| % Urban | 25<sup>th</sup> | 25.1 | 80<sup>th</sup> | 65.1 | 95<sup>th</sup> | 87.5 |
| % Access to Water Systems | 25<sup>th</sup> | 55.3 | 50<sup>th</sup> | 74.4 | 75<sup>th</sup> | 87.1 |
| % Inadequate Housing Conditions | 25<sup>th</sup> | 21.4 | 50<sup>th</sup> | 40.1 | 75<sup>th</sup> | 54.8 |
| % Living in Low-income Areas | 10<sup>th</sup> | 79.2 | 25<sup>th</sup> | 91.9% | 75<sup>th</sup> | 99.4% |

**Table 3.** Final models for temperature and rainfall conditions.

| Description | Model Formula | DIC | CV Mean Log Score |
| --- | --- | --- | --- |
| Baseline<sub>Reg</sub> | $\alpha + \beta_{s(s)} m(t) + \varphi_{s a(t)} + v_{s a(t)}$ | 519404 | 1.712 |
| Baseline<sub>Reg</sub> + MaxIntensity<sub>Warmspell</sub> *Elev | $\alpha + \beta_{s(s)} m(t) + \varphi_{s a(t)} + v_{s a(t)} + \gamma_r + f.w(x_{1st}, l) * v_{ELEV}$ | 490792 | 1.572 |
| Baseline<sub>Reg</sub> + WetDays *Urb | $\alpha + \beta_{s(s)} m(t) + \varphi_{s a(t)} + v_{s a(t)} + \gamma_r + f.w(x_{1st}, l) * v_{URB}$ | 511635 | 1.663 |
| Baseline<sub>Reg</sub> + DryDays *Urb + MaxIntensity<sub>Warmspell</sub> *Elev | $\alpha + \beta_{s(s)} m(t) + \varphi_{s a(t)} + v_{s a(t)} + \gamma_r + f.w(x_{1st}, l) * v_{ELEV} + f.w(x_{2st}, l) * v_{URB}$ | 494009 | 1.431 |

**Table 4.** Maximum relative risk across elevation and percent urban gradients.

|  | Low % Urban<br>25% | Med. % Urban<br>40% | High % Urban<br>60% |
| --- | --- | --- | --- |
| **High Elevation** | 3.39 [1.70, 6.75] | 3.40 [1.83, 5.21] | 3.42 [1.24, 7.88] |
| **Med Elevation** | 3.93 [2.11, 7.10] | 3.94 [1.64, 6.34] | 3.93 [2.57, 7.64] |
| **Low Elevation** | 4.48 [2.46, 5.96] | 2.12 [1.19, 5.10] | 4.49 [3.20, 6.52] |

We selected a climate extremes-urban characteristics interaction model for each of the three climate conditions (temperature, dry conditions, and wet conditions) by comparing models of increasing complexity. For each, we calculated goodness of fit measures, including deviance information criterion (DIC), which balances model accuracy against complexity by penalizing for the number of effective parameters in the model, and the mean cross-validated log score, which measures the predictive power of the model when excluding one data point at a time^54,55^. With the DIC and log score, smaller values indicate better-fitting models. We also calculated the difference in mean absolute error (MAE) between the climate extremes models compared to the baseline models as well as to the standard weather variable models (e.g., T_mean_, Total-Precip). This provided us with an understanding of whether the climate extremes models better predicted dengue cases compared to baseline or standard weather variable models; and where this added value was geographically.

We constructed a final model by combining the best-fitting extreme temperature and rainfall conditions models (with elevation and urbanicity interactions). With this model, we quantified the lagged and non-linear effects of climate extremes on dengue risk across elevation and urbanization gradients. Finally, we cross-validated our predictions, by refitting the selected model 12×12 times, excluding a month per year from the fitting process each time. We compared observations from each department to their respective out-of-sample posterior predictive distributions between January 2008 and December 2019.

## Data Availability

All data produced in the present work are contained in the manuscript

## Acknowledgements

We thank Xandre Chourio and Ángel Muñoz (Columbia University) for support with accessing ENACTS climate data through the International Research Institute for Climate and Society (IRI). PK was supported in part by the NSF Geography and Spatial Sciences—Doctoral Dissertation Improvement Grant. RL was supported by the Wellcome Trust funded project IDExtremes (226069/Z/22/Z) and a Royal Society Dorothy Hodgkin Fellowship; as well as E4Warning, financed by the European Union’s Horizon Europe research and innovation programme (101086640).

