## Supplementary material for "Climate extremes increase dengue risk along elevation and socio-economic gradients in Colombia"

**Colombia**

**1. Supplementary Methods**

A. Study Area

Colombia is located in South America’s northwest corner, between the Pacific Ocean and Caribbean Sea. The country is geographically and topographically complex, with the Andes Mountains dominating the central portion of the country, reaching up to 5,400 meters in elevation and making up over 287,000 km^2^ in land area^1^. The Andes consist of three primary ranges (the Western, Central, and Eastern Cordilleras), separated by the Cauca and Magdalena River Valleys; peripheral mountain ranges add to the landscape complexity. Additionally, the Amazon Rainforest is located in the south and extends to the country’s southern and southwestern borders. The country is partitioned into five regions—the Pacific, Caribbean, Andean, Orinoco, and Amazon (Fig. S1a). Colombia’s mainland (excluding the islands of San Andrés and Providencia) includes 31 departments and 1,120 municipalities, with an estimated population of 48.3 million people (2018) (Fig. S1c)^2^. Nearly 80% of Colombia’s residents live in urban settlements, spanning a range of population sizes and elevations, however the majority of the country’s population lives in the Andean region (Fig. S1a, Fig. S2)^2^.

With respect to socio-economic conditions, in recent decades there have been increased national investments in the water and sanitation sectors, providing resources to municipal governments and a level of infrastructure standardization across the country^3^. In contrast, the housing sector receives little national-level funding and dwelling conditions are determined primarily by municipality-level regulations and land and housing market conditions^3^. This contributes to a high degree of spatial variation and disparity in whether or not the populations’ housing needs are met (Fig. S3). Colombian cities have high rates of informal settlement, with high population density and building densities, in which low-income neighborhoods are often built in environmentally-hazardous zones^4^. Finally, challenges in constructing road networks across mountainous terrain have meant that, historically, the country has had limited connective infrastructure, with greater emphasis on regional trade and movement^3^.

B. Climatology

Inter-annual variation and extreme climatic events (e.g. warm spells, dry spells) in Colombia are driven by atmospheric-oceanic interactions, including the El Niño Southern Oscillation (ENSO). While El Niño produces warmer and drier conditions compared to mean conditions, La Niña is associated with wetter conditions—particularly in the central, northern, and western regions of Colombia. El Niño events were observed during 2009–2010, 2015–2016, and 2018–2019, with 2015–2016 being among the strongest ENSO events on record. La Niña years were observed during 2007–2008, 2010–2011, 2012, and 2017–2018^5,6^ .

The primary process driving spatial variation in intra-annual rainfall variation is the latitudinal position of the Intertropical Convergence Zone (ICTZ), as well as the interaction of the ICTZ with the Andes, the Atlantic and Pacific Oceans, and the Amazon rainforest^7^. These localized phenomena contribute to the five regions having unique climates, characterized by distinct rainfall patterns (Fig. S1b)^8,9^. An analysis by Urrea et al. characterized precipitation in Colombia by four regimes. A unimodal regime, with one rainy and one dry season, is observed in parts of the Orinoco, Amazon, and Caribbean regions. Specifically, the Orinoco is characterized by a short rainy season during May–July; and while the Caribbean region receives less rainfall overall compared to other areas (1,070 mm annually), it has an extended rainy period during April–November, when rainfall tends to be greater than 100 mm per month. Overall, the Andes has a bimodal annual rainfall regime. Bimodal regimes are observed due to the mountainous environment and latitudinal displacement of the ITCZ, such that during December-January-February, the ITCZ is south of the equator, bringing moisture to latitudes south of 2°N. During March-April-May, the ITCZ shifts north (2°–7°N); while during June-July-August the ITCZ moves even further north (reducing rainfall in the Andes). Finally, during September-October-November, the ITCZ moves southward again (2°–7°N), resulting in bimodal rainfall, particularly for the departments of Antioquia, Caldas, Cundinamarca, Quindío, Risaralda, and Tolima. Transition zones, between regions, tend to experience a blended regime, with a combination of unimodal and bimodal patterns^10^.

These factors contribute to four primary climatic zones. Low-elevation zone (<900 m), which have alternating rainy and dry seasons. Temperate zones (900–1980 m), which includes two wet seasons during January–March and July–September. Cold zones (1980–3500 m), which have two wet seasons during April–May and September–December. Finally, at the highest elevation are the Paramos (3500–4500 m), which extend to the “permanent” snowline^11^.

C. Data Sources

*Dengue Cases*

We obtained data on dengue fever cases (including both classic dengue fever and dengue hemorrhagic fever) from the national surveillance system, SIVIGILA (*Sistema Nacional de Vigilancia en Salud Pública*), administered by the National Institute of Health (*Instituto Nacional de Salud*). In Colombia, dengue fever is a notifiable disease, therefore all probable and confirmed cases must be reported from municipalities to the national level by law. Cases of classic dengue fever (without warning signs) (SIVIGILA Code: 210) are reported weekly, whereas cases of dengue hemorrhagic fever (Code: 220) are reported immediately^12^. Probable cases are based on: 1) clinical diagnosis with two or more symptoms and at least one serological immunoglobulin M (IgM) test; OR 2) an epidemiological link to a confirmed case within 14 days of symptom onset. Confirmed cases of dengue are based on laboratory diagnosis via: 1) PCR within five days of symptom onset; OR 2) IgM serology with Dengue ELISA kits after 5 days of symptom onset. During an outbreak, serological samples are taken from 5% of cases of classic dengue fever and for all cases of dengue hemorrhagic fever. Case definitions of dengue were changed in January 2010, as new WHO definitions of dengue fever were adopted.

National surveillance data for Colombia has several considerations, including the spatial and temporal scales of the available data and analysis. Monthly-level case aggregation precluded examining short-term effects of extreme climatic events on dengue risk. However, weekly-level case data were sparse across many municipalities. Previous investigations have revealed reporting lags in national surveillance that are partially relieved when aggregating case data to monthly timescales. However, under-reporting persists, particularly for mild cases of classic dengue fever^13,14^. Evaluations of SIVIGILA have demonstrated that confirmed dengue case counts taken from department-level health agencies are 5.8 times higher than national-level data using data from an active surveillance program^13,15^. This may be because under-resourced jurisdictions have limited healthcare infrastructure or laboratory capacity, which prevent the use of national dengue case definitions or result in misdiagnoses. Additionally, under-reporting may reflect at-home care for mild febrile illnesses^16^. Reporting biases may also vary based on epidemiological context—where increased public awareness of dengue risk during outbreaks reduces under-reporting rates. Finally, through SIVIGILA, data on dengue serotypes were not available.

*Demographic and Socio-economic Data*

We collated population data from Colombia’s National Administrative Department of Statistics (*Departamento Administrativo Nacional de Estadística*, DANE). Municipality population data for 2007–2017 were taken from population projections using the 2005 national census; data for 2018–2019 were taken from population estimates and projections using the 2018 census^2,17^. Socio-economic data were also available for each municipality using the 2018 census. For this study we considered information on: access to public services (percent with access to: electricity, piped water, sewage systems, trash collection, and internet), housing types, and household-level population density (average number of: households per building, individuals per room)^2^.

*Climate*

We obtained daily mean temperature from Colombia ENACTS initiative of the International Research Institute for Climate and Society^18^. The dataset provides daily temperature from 1980–2019 at 0.1° spatial resolution (~11.1 km). Data from Colombia ENACTS are blended (i.e., spatially and temporally interpolated and smoothed) using NASA’s MERRA-2 data and ground-truth records gathered at weather stations operated by the Colombian Institute of Hydrology, Meteorology and Environmental Studies (*Instituto de Hidrología, Meteorología y Estudios Ambientales*, IDEAM)^19^. Daily mean temperature values were then used to derive monthly temperature variables (Table S1). We obtained daily precipitation data from the Climate Hazards Group Infrared Precipitation with Station data (CHIRPS)^20^. CHIRPS provides daily precipitation estimates from 1981–2019 at 0.05° spatial resolution^20^. At the tropics, the 0.05° spatial resolution equates to ~5 km, resulting in 37,012 CHIRPS grid cells across Colombia. Funk et al. previously validated CHRIPS for use in Colombia using IDEAM precipitation stations^20^. Results indicated CHIRPS data are well correlated with station data, although product biases exist (particularly for rainy seasons, when errors are associated with an overestimation of daily precipitation values)^20,21^.

D. Model Description

*Spatiotemporal Bayesian hierarchical model*

We constructed a spatiotemporal Bayesian hierarchical mixed effects model, with a response variable of monthly counts of dengue cases for 1,120 Colombian municipalities during January 2008 to December 2019. We assumed that monthly dengue cases, $y_{st}$ (*s* = 1, …, 1,120; *t* = 1, …, 132) followed a negative binomial distribution:

$$y_{st} | \mu_{st}, \kappa\sim NegBin(\mu_{st}, \kappa)$$

$$log(\mu_{st}) = log(p_{sa(t)}) + log(\rho_{st})$$

where $\mu_{st}$was the distribution mean equal to the annual population per 100,000 $p_{sa(t)}$, where $a(t)$ = 2008, …, 2019, multiplied by the unknown dengue incidence rate estimate $\rho_{st}$ for municipality *s* at month *t*, with κ representing the overdispersion parameter. We accounted for population effects using an offset parameter of log population per 100,000 at the linear predictor scale (coefficient of one), so that $\rho_{st}$ was equivalent to the incidence rate per 100,000 population.

To estimate model parameters we used Integrated Nested Laplace Approximation (INLA, www.r-inla.org) in R. We accounted for parameter uncertainty by assigning prior distributions to the parameters. With R-INLA, we established a baseline model with spatiotemporal random effects to account for seasonal variability at the department level, and interannual variability at the municipality level with spatial dependency structures. For all random effects, we used the Penalized Complexity (PC) priors for the precision $t$ = 1/$\sigma^{2}$ , so that Pr(1/√t > 0.5) = 0.01.

Department-level random effects were included per calendar month, $m\left( t \right)$= 1, …, 12 (January to December), for each department, $d\left( s \right)$ = 1, …, 31, to account for differences in vector ecology, health care instrastructure and access, as well as dengue surveillance. We assigned this department-specific monthly random effect, $\beta_{d(s) m(t)}$, a cyclic random walk of order one (i.e., first difference prior distribution) so that each effect is derived from the immediately preceding effect,

$$\beta_{d(s) m(t)} - \beta_{d\left( s \right)m\left( t \right)-1}\sim N(0, {\sigma^{2}}_{\beta})$$

, where $\beta_{d(s)1}$ represents the parameter estimate for January for department $d\left( s \right)$. The cyclic nature of this prior distribution allowed each month to depend on the previous month, with no discontinuity between January in year $a(t)$ and December of the previous year, $a(t)$-1.

Separately, we included year-specific random effects at the municipality level to account for annual variation and long-term trends, interacting both spatially-unstructured and spatially-structured components. For this, we used a modified Besag-York-Mollie (BYM) model with a scaled spatial component, which assigns hyperpriors that are transferable between geographic settings. The modified BYM model consists of a precision parameter and a mixing parameter. The precision parameter represents the marginal precision and controls variability explained by the spatial effect. While the mixing parameter distributes the variability between spatially-unstructured and spatially-structured components, $\varphi_{s a(t)}+\upsilon_{s a(t)}.$The spatially-unstructured component, $\varphi_{s a(t)}$, accounts for unobserved confounding factors between municipalities, including population-level immunity, healthcare access and services, etc. The spatially-structured component, $\upsilon_{s a(t)}$, assumes that spatial dependency exists between neighboring municipalities, accounting for spatial autocorrelation. Finally, we tested whether including the five bioclimatic regions and six Lang climate classifications as fixed effects, $\gamma_{r}$ and $\gamma_{c}$ , respectively, improved the model fit.

*Distributed lag nonlinear model (DLNM)*

We used DLNMs to explore nonlinear and lagged associations between dengue incidence rates and climate covariates. The DLNM is rooted in a cross-basis function, which is a bi-dimensional space of functions specifying dependency across the space of both the predictor and lags. Cross-basis functions are constructed by combining an exposure–response function, f(x), with a lag-response function, w(l), resulting in f.w(x, l). We assessed the delayed effects of climate covariates for up to six months, using natural cubic splines for both the exposure (two equally-spaced knots) and the lag (one internal knot at the 50th percentile for temperature covariates; two equally spaced knots for precipitation covariates).

The number of coefficients in the cross-basis matrix represents the degrees of freedom (df) used to model the association. Temperature DLNMs were defined by nine cross-basis variables (3 df [natural cubic spline with 1 knot + 1] in the exposure-response dimension multiplied by 3 df [natural cubic spline with 1 knot + intercept + 1] in the lag-response dimension). Precipitation DLNMs were defined by twelve cross-basis variables (3 df [natural cubic spline with 2 knots + 1] in the exposure-response dimension multiplied by 4 df [natural cubic spline with 2 knots + intercept + 1] in the lag-response dimension).

To assess the impact of extreme temperature conditions along an elevation gradient, we tested the linear interaction between extreme temperature variables and the minimum elevation of the urban extent for each municipality^22^. We centered minimum elevation values at 0, 1,000, and 1,750 meters to partition low-, intermediate-, and high-elevation municipalities. By centering, or subtracting these key values from the given municipality value, we parameterized the interaction term (the cross-basis multiplied by the interaction variable, i.e., minimum elevation) as 0 for a given partition. Therefore predictions given by the main term (the standard cross-basis) are interpreted as the exposure-lag-response for a given partition only. By varying this centering point, we held the model constant, but changed the parameterization.

For dry conditions, we used linear interaction terms by centering continuous variables at low values (between the 10^th^–25^th^ percentile, depending on the variable), intermediate (25^th^–50^th^ percentile), and high (75^th^ percentile) values for each variable (Main Text, Table 2). At the highest level of granularity for socio-economic condition, we tested the percent of the population in an urban area. We then examined the municipality's water infrastructure level, measured by the percentage of residents accessing water systems. And at the lowest level of granularity, we tested whether household-level properties may reveal the need for water storage, using a summary index calculated from the 2018 Colombian national census measuring household conditions (which includes survey questions on public services and the structural stability of the dwelling). For excess rainfall conditions, we aimed to capture the interaction between heavy rainfall and the availability of water-holding containers in the urban landscape. We used linear interaction terms for socio-economic characteristics, including the percent urban, population density, and the percent of the population living in low socio-economic status neighborhoods. We centered these continuous variables at low, intermediate, and high values (Main Text, Table 2).

We calculated goodness of fit measures to select the best-fitting model, including the deviance information criterion (DIC) and the cross validated (CV) log score. We also calculated the difference in mean absolute error (MAE) between the baseline model and each new candidate model, to identify the proportion of municipalities for which more complex models improved model fit. The best estimate of the dengue incidence rate was given with the following model formulas:

$$y_{st} | \mu_{st}, \kappa\sim NegBin(\mu_{st}, \kappa)$$

$$log(\mu_{st}) = log(p_{sa(t)}) + log(\rho_{st})$$

$$log\left( p_{sa\left( t \right)} \right)= \alpha+\beta_{d(s) m(t)}+\varphi_{s a(t)}+\upsilon_{s a(t)}+\gamma_{r}+ f.w\left( x_{1st}, l \right)+ f.w\left( x_{1st}, l \right)*v_{ELEV}+f.w\left( x_{2st}, l \right)+f.w(x_{2st}, l)*v_{URB}$$

**2. Supplementary Tables**

Table S1. Principal component (PC) factor loadings for socio-economic variables and elevation for Colombian municipalities

| **Variables** | **PC1** | **PC2** | **PC3** |
| --- | --- | --- | --- |
| Population Size | -0.14 | 0.11 | -0.7 |
| % Living in Urban Areas | -0.37 | 0.35 | 0.13 |
| Population Density | -0.21 | 0.25 | -0.45 |
| Minimum elevation (m) | -0.09 | -0.62 | -0.16 |
| % Low-income Housing | 0.31 | 0 | 0.32 |
| % Access to Trash Collection | 0.42 | -0.2 | -0.15 |
| % Access to Water Systems | 0.35 | 0.09 | -0.32 |
| % Access to Sewage Treatment | 0.43 | -0.05 | -0.16 |
| % Housing Deficiencies | 0.41 | 0.28 | -0.08 |
| % Overcrowded Housing | 0.19 | 0.53 | 0.11 |

Table S2. Models of extreme temperature conditions with increasing complexity

| **Description** | **Model Formula** | **DIC** | **CV Mean Log Score** |
| --- | --- | --- | --- |
| Baseline | α + β_s(s) m(t)_ + φ_s_ _a(t)_ + υ_s a(t)_ | 519404 | 1.712 |
| Baseline_ClimZone_ | α + β_s(s) m(t)_ + φ_s_ _a(t)_ + υ_s a(t)_ + γ_c_ | 518307 | 1.706 |
| Baseline_Reg_ | α + β_s(s) m(t)_ + φ_s_ _a(t)_ + υ_s a(t)_ + γ_r_ | 517182 | 1.695 |
| Baseline_Reg_ + T_mean_ | α + β_s(s) m(t)_ + φ_s_ _a(t)_ + υ_s a(t)_ + γ_r_ + f.w(x_1st_, *l*) | 501768 | 1.628 |
| Baseline_Reg_ + WarmDays | α + β_s(s) m(t)_ + φ_s_ _a(t)_ + υ_s a(t)_ + γ_r_ + f.w(x_1st_, *l*) | 516194 | 1.681 |
| Baseline_Reg_ + MaxDuration_Warmspell_-T_90_ | α + β_s(s) m(t)_ + φ_s_ _a(t)_ + υ_s a(t)_ + γ_r_ + f.w(x_1st_, *l*) | 516161 | 1.681 |
| Baseline_Reg_ + MaxIntensity_Warmspell_ | α + β_s(s) m(t)_ + φ_s_ _a(t)_ + υ_s a(t)_ + γ_r_ + f.w(x_1st_, *l*) | 516101 | 1.680 |
| Baseline_Reg_ + MaxIntensity_Warmspell_ + Elev | α + β_s(s) m(t)_ + φ_s a(t)_ + υ_s a(t)_ + γ_r_ + f.w(x_1st_, *l*) + v_ELEV_ | 490928 | 1.572 |
| Baseline_Reg_ + MaxIntensity_Warmspell_ *Elev | α + β_s(s) m(t)_ + φ_s a(t)_ + υ_s a(t)_ + γ_r_ + f.w(x_1st_, *l*)*v_ELEV_ | 490792 | 1.572 |

Table S3. Models of dry conditions with increasing complexity

| **Description** | **Model Formula** | **DIC** | **CV Mean Log Score** |
| --- | --- | --- | --- |
| Baseline_Reg_ | α + β_s(s) m(t)_ + φ_s_ _a(t)_ + υ_s a(t)_ + γ_r_ | 519404 | 1.712 |
| Baseline_Reg_ + Total-Precip. | α + β_s(s) m(t)_ + φ_s_ _a(t)_ + υ_s a(t)_ + γ_r_ + f.w(x_1st_, *l*) | 516895 | 1.695 |
| Baseline_Reg_ + MaxDuration_DrySpell_ | α + β_s(s) m(t)_ + φ_s_ _a(t)_ + υ_s a(t)_ + γ_r_ + f.w(x_1st_, *l*) | 516750 | 1.693 |
| Baseline_Reg_ + DryDays | α + β_s(s) m(t)_ + φ_s_ _a(t)_ + υ_s a(t)_ + γ_r_ + f.w(x_1st_, *l*) | 516594 | 1.694 |
| Baseline_Reg_ + DryDays + Urb | α + β_s(s) m(t)_ + φ_s a(t)_ + υ_s a(t)_ + γ_r_ + f.w(x_1st_, *l*) + v_URB_ | 511658 | 1.664 |
| Baseline_Reg_ + DryDays + WaterSys | α + β_s(s) m(t)_ + φ_s a(t)_ + υ_s a(t)_ + γ_r_ + f.w(x_1st_, *l*) + v_WAT_ | 512592 | 1.670 |
| Baseline_Reg_ + DryDays + Housing | α + β_s(s) m(t)_ + φ_s a(t)_ + υ_s a(t)_ + γ_r_ + f.w(x_1st_, *l*) + v_HOUS_ | 514072 | 1.677 |
| Baseline_Reg_ + DryDays *Urb | α + β_s(s) m(t)_ + φ_s a(t)_ + υ_s a(t)_ + γ_r_ + f.w(x_1st_, *l*)*v_URB_ | 511638 | 1.663 |

Table S4. Models of wet conditions with increasing complexity

| **Description** | **Model Formula** | **DIC** | **CV Mean Log Score** |
| --- | --- | --- | --- |
| Baseline_Reg_ | α + β_s(s) m(t)_ + φ_s_ _a(t)_ + υ_s a(t)_ + γ_r_ | 519404 | 1.712 |
| Baseline_Reg_ + Total-Precip | α + β_s(s) m(t)_ + φ_s_ _a(t)_ + υ_s a(t)_ + γ_r_ + f.w(x_1st_, *l*) | 516895 | 1.695 |
| Baseline_Reg_ + Anomaly_Precip_ | α + β_s(s) m(t)_ + φ_s_ _a(t)_ + υ_s a(t)_ + γ_r_ + f.w(x_1st_, *l*) | 517472 | 1.695 |
| Baseline_Reg_ + MaxDuration_WetSpell_ | α + β_s(s) m(t)_ + φ_s_ _a(t)_ + υ_s a(t)_ + γ_r_ + f.w(x_1st_, *l*) | 517189 | 1.695 |
| Baseline_Reg_ + WetDays | α + β_s(s) m(t)_ + φ_s_ _a(t)_ + υ_s a(t)_ + γ_r_ + f.w(x_1st_, *l*) | 516781 | 1.692 |
| Baseline_Reg_ + WetDays + Urb | α + β_s(s) m(t)_ + φ_s a(t)_ + υ_s a(t)_ + γ_r_ + f.w(x_1st_, *l*) + v_URB_ | 511668 | 1.663 |
| Baseline_Reg_ + WetDays + PopDen | α + β_s(s) m(t)_ + φ_s_ _a(t)_ + υ_s a(t)_ + γ_r_ + f.w(x_1st_, *l*) + v_POP_ | 512591 | 1.670 |
| Baseline_Reg_ + WetDays + LowSES | α + β_s(s) m(t)_ + φ_s_ _a(t)_ + υ_s a(t)_ + γ_r_ + f.w(x_1st_, l) + v_SES_ | 516611 | 1.691 |
| Baseline_Reg_ + WetDays *Urb | α + β_s(s) m(t)_ + φ_s a(t)_ + υ_s a(t)_ + γ_r_ + f.w(x_1st_, *l*)*v_URB_ | 511635 | 1.663 |

Table S5. Extreme Temperature Models Compared to Mean Temperature Model

| **Region** | **Added Value** | **Total** | **Proportion** |
| --- | --- | --- | --- |
| **Comparison: Days-T90 vs. Tmean** | | | |
| Amazon | 0 | 47 | 0% |
| Andean | 212 | 767 | 27.6% |
| Caribbean | 49 | 206 | 23.8% |
| Orinoco | 4 | 56 | 7.1% |
| Pacific | 13 | 44 | 29.5% |
| **Comparison: Days-T90 vs. Tmean** | | | |
| Amazon | 0 | 47 | 0% |
| Andean | 214 | 767 | 27.9% |
| Caribbean | 53 | 206 | 25.7% |
| Orinoco | 4 | 56 | 7.1% |
| Pacific | 14 | 44 | 31.8% |
| **Comparison: Max-Anomaly vs. Tmean** | | | |
| Amazon | 2 | 47 | 2.3% |
| Andean | 218 | 767 | 28.4% |
| Caribbean | 51 | 206 | 24.8% |
| Orinoco | 5 | 56 | 8.9% |
| Pacific | 15 | 44 | 34.1% |
| **Comparison: Max-Anomaly + Elevation vs. Tmean** | | | |
| Amazon | 24 | 47 | 51.1% |
| Andean | 407 | 767 | 53.1% |
| Caribbean | 101 | 206 | 49.0% |
| Orinoco | 20 | 56 | 35.7% |
| Pacific | 30 | 44 | 68.2% |
| **Comparison: Max-Anomaly*Elevation vs. Tmean** | | | |
| Amazon | 25 | 47 | 51.1% |
| Andean | 404 | 767 | 52.7% |
| Caribbean | 102 | 206 | 49.4% |
| Orinoco | 21 | 56 | 37.5% |
| Pacific | 32 | 44 | 72.2% |

**3. Supplementary Figures**

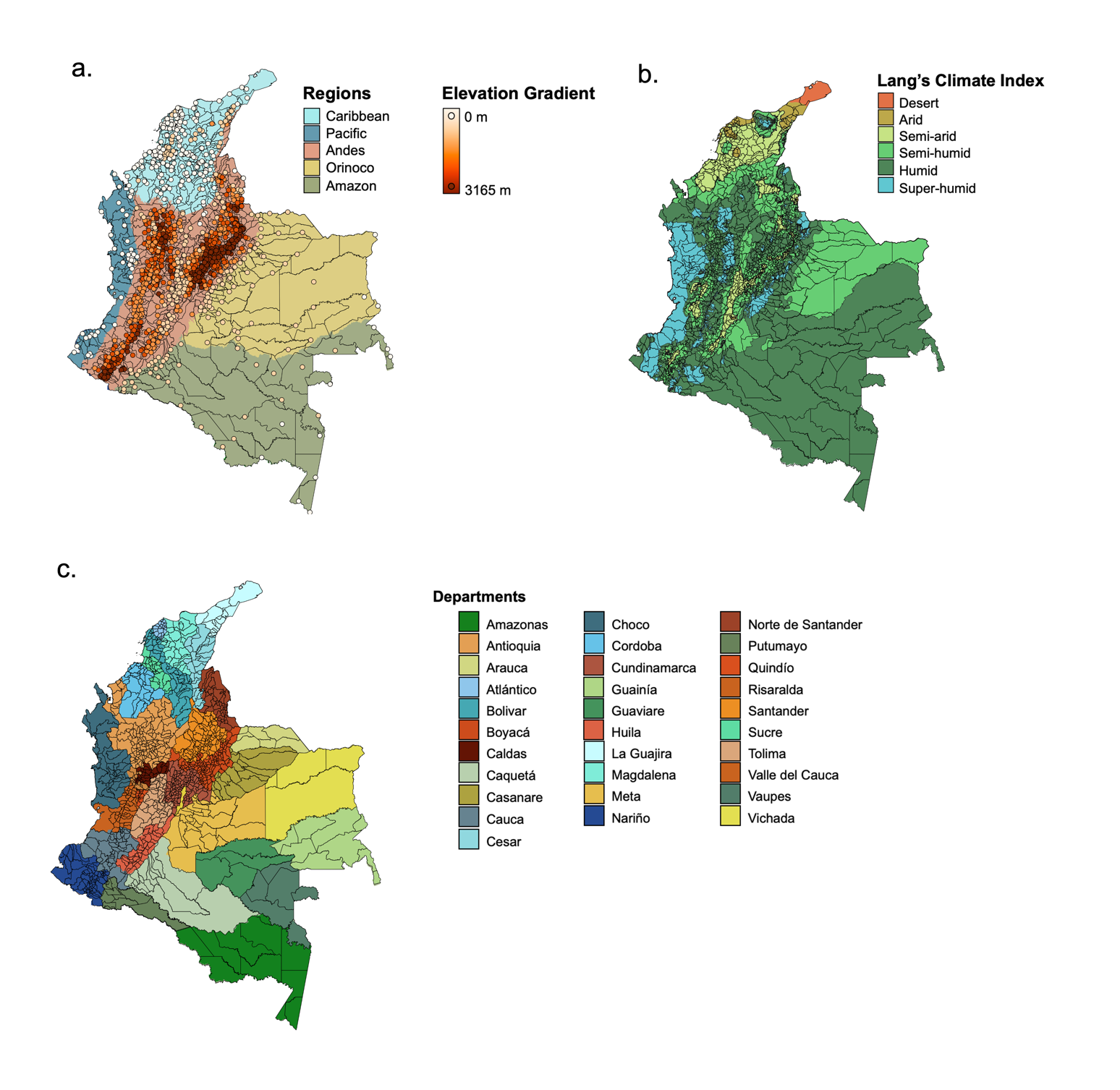

**Fig. S1. Bioclimatic and political regions of Colombia.**

**a.** The five bioclimatic regions of Colombia with the centroids of the urban centers for 1120 mainland municipalities, shaded according to minimum elevation of the urban extent. Data sources: IDEAM, SRTM^23^. **b.** The six Lang climate index classifications for Colombia. Data source: IDEAM. **c.** The 31 departments of Colombia. Data source: DANE.

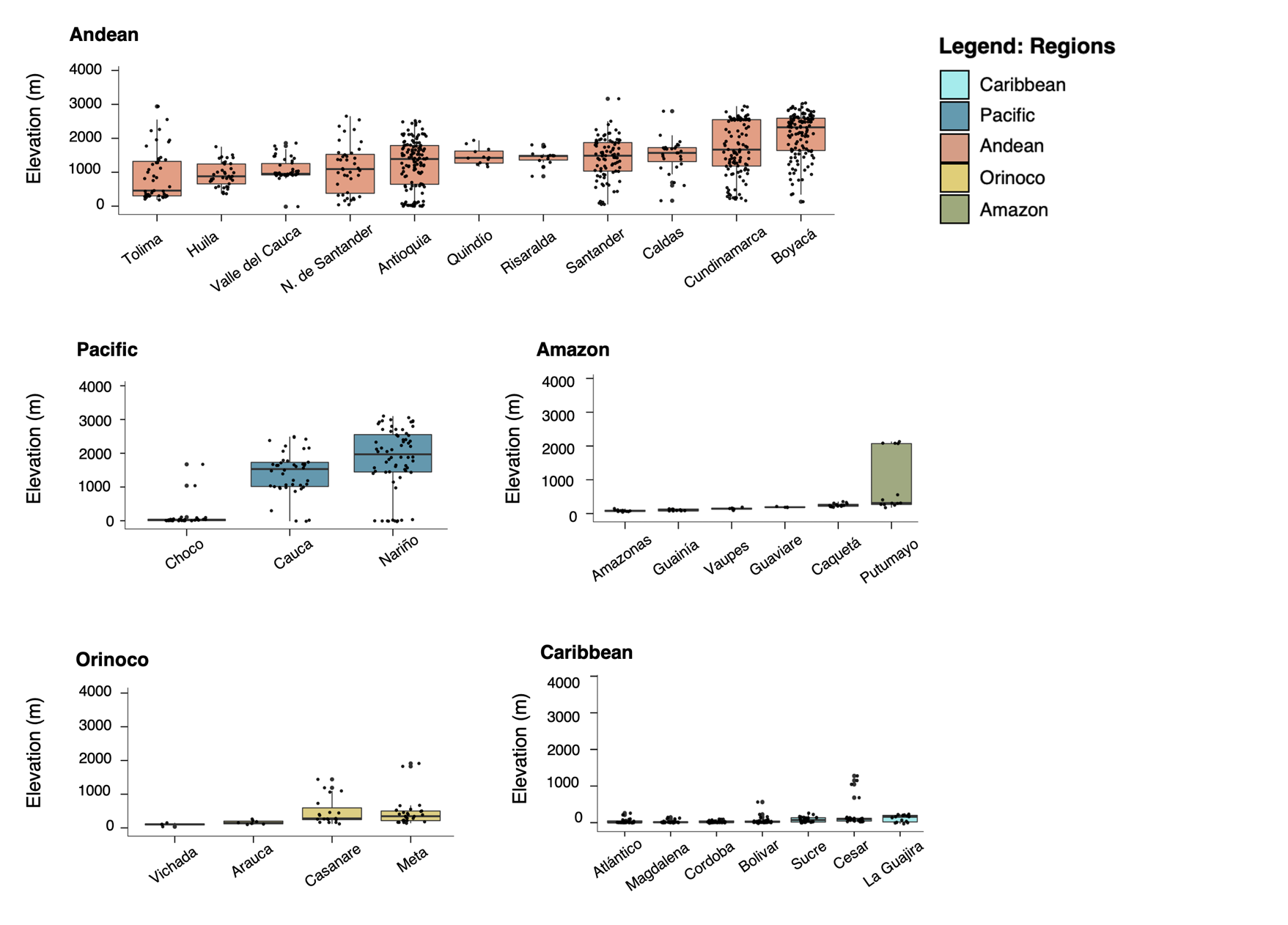

**Fig. S2. Elevation ranges per Colombian Department.**

Boxplot of elevation ranges within each Colombian department. Individual points represent the minimum elevation of the urban extent for a municipality within the department. Departments are grouped and colored according to bioclimatic region. Data source: SRTM^23^.

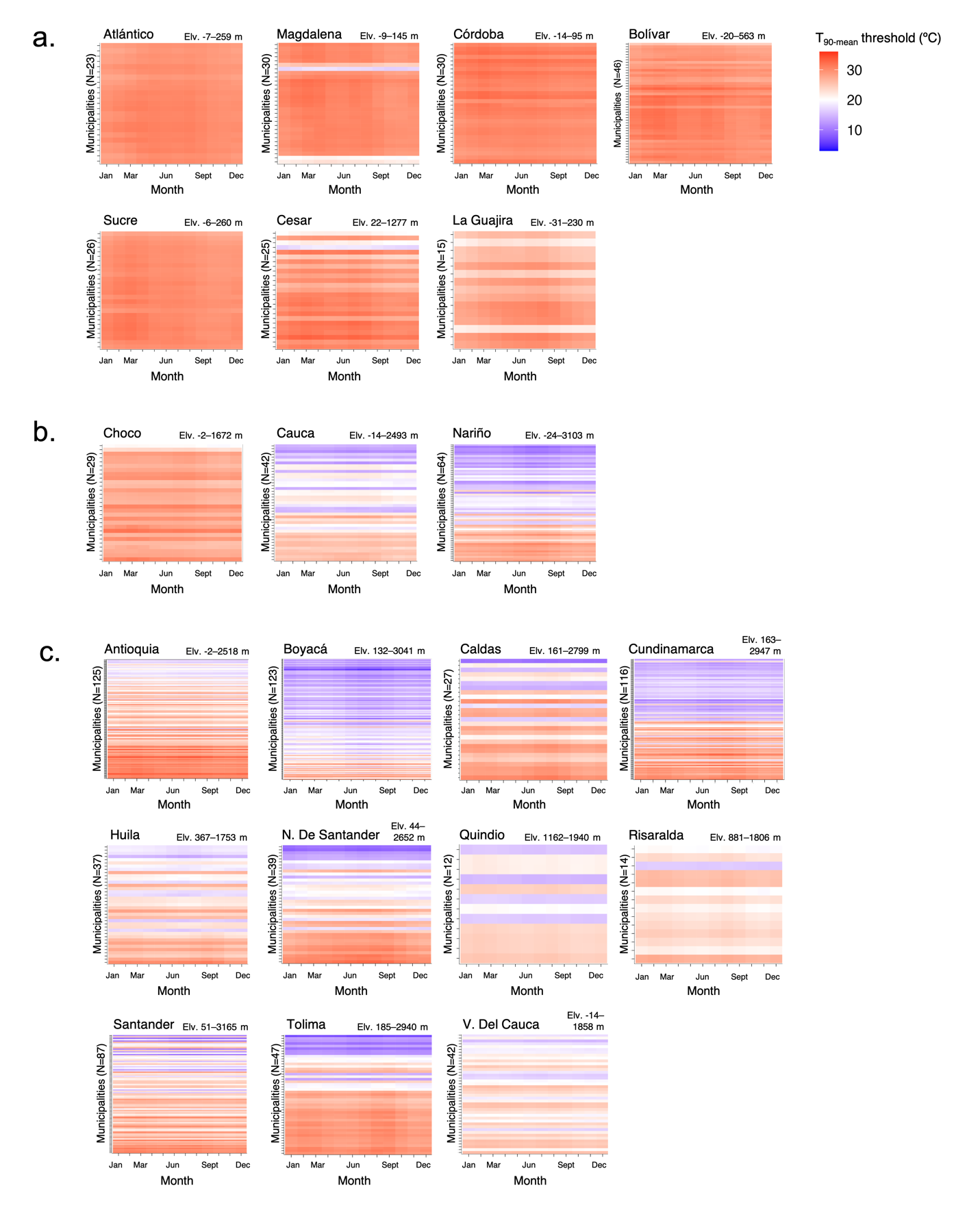

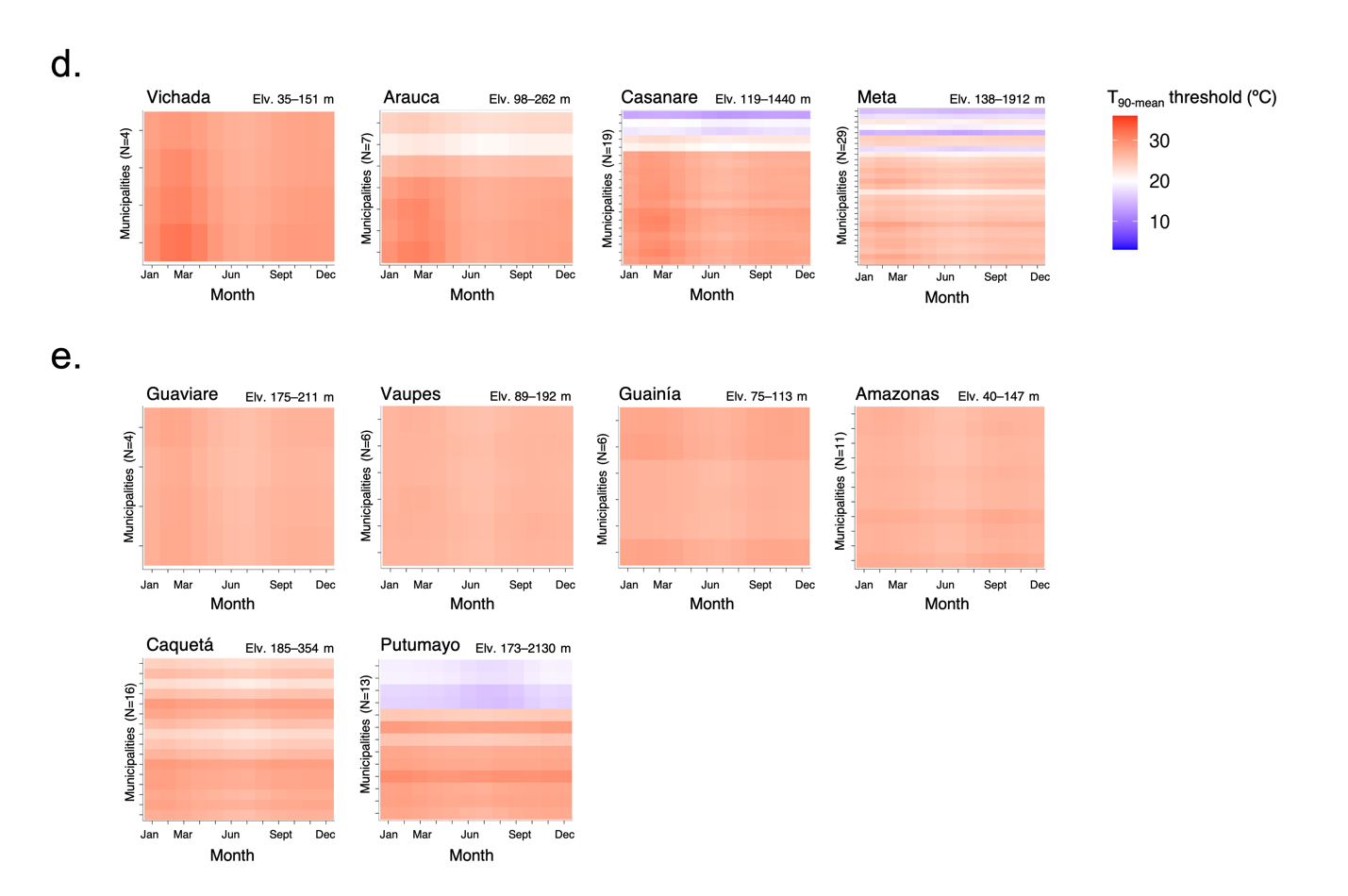

**Fig. S3. Heatmap of the municipality monthly T_90-mean_ threshold grouped by department and region.**

Municipality monthly T_90-mean_ threshold from January to December for the 31 mainland departments in Colombia. with municipalities listed on the y-axis in descending order by minimum elevation of the municipality’s urban extent. The range of elevation values in each department is provided. Departments are grouped according to their bioclimatic region. **a.** For the Caribbean region, we see primarily low-elevation municipalities and higher T_90-mean_ threshold values between 27–31°C, with minimal intra-annual variation. **b.** For the Pacific, we observe T_90-mean_ threshold values that varied according to elevation, with minimal intra-annual variation. **c.** For the Andes, we observe T_90-mean_ threshold values that varied according to elevation, with minimal intra-annual variation. **d.** For the Orinoco, we observe T_90-mean_ threshold values that varied according to elevation. Values are slightly higher for January–April, compared to the rest of the year. **e.** For the Amazon, we observe primarily low-elevation municipalities, with monthly T_90-mean_ thresholds primarily ranging between 25–26°C, with the exception of four high-elevation municipalities in Putumayo.

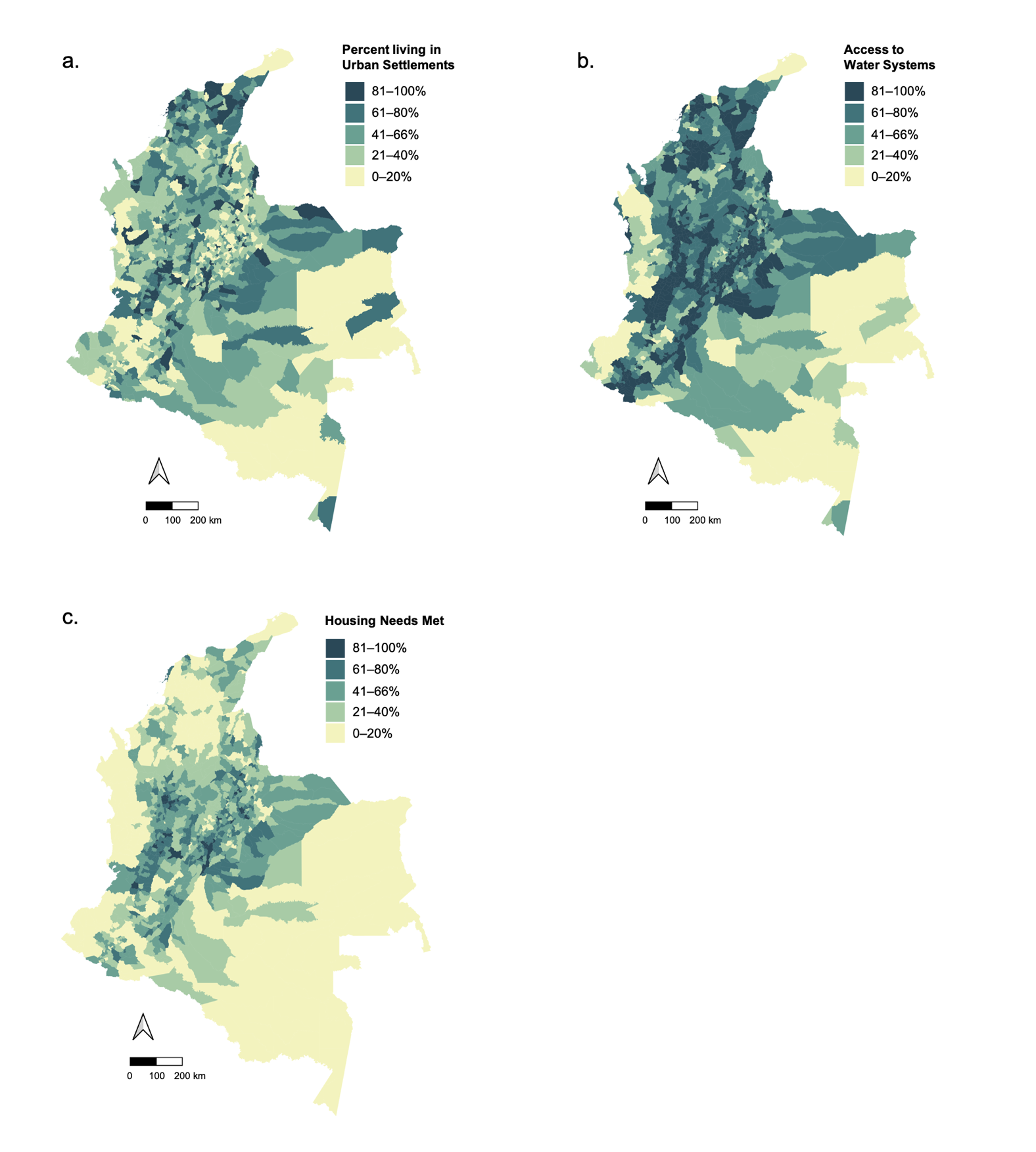

**Fig. S4. Socio-economic conditions at the municipality level in Colombia.**

**a**. The percent of the municipality population living in an urban area (i.e., *cabecera municipal*)^2^. **b.** The percent of the municipality population with access to water systems, either public or community-based (i.e., *la cobertura de servicios de acueductos*). **c.** The percent of homes within the municipality with basic housing needs met (e.g., access to public services, no structural deficiencies) ^24^.

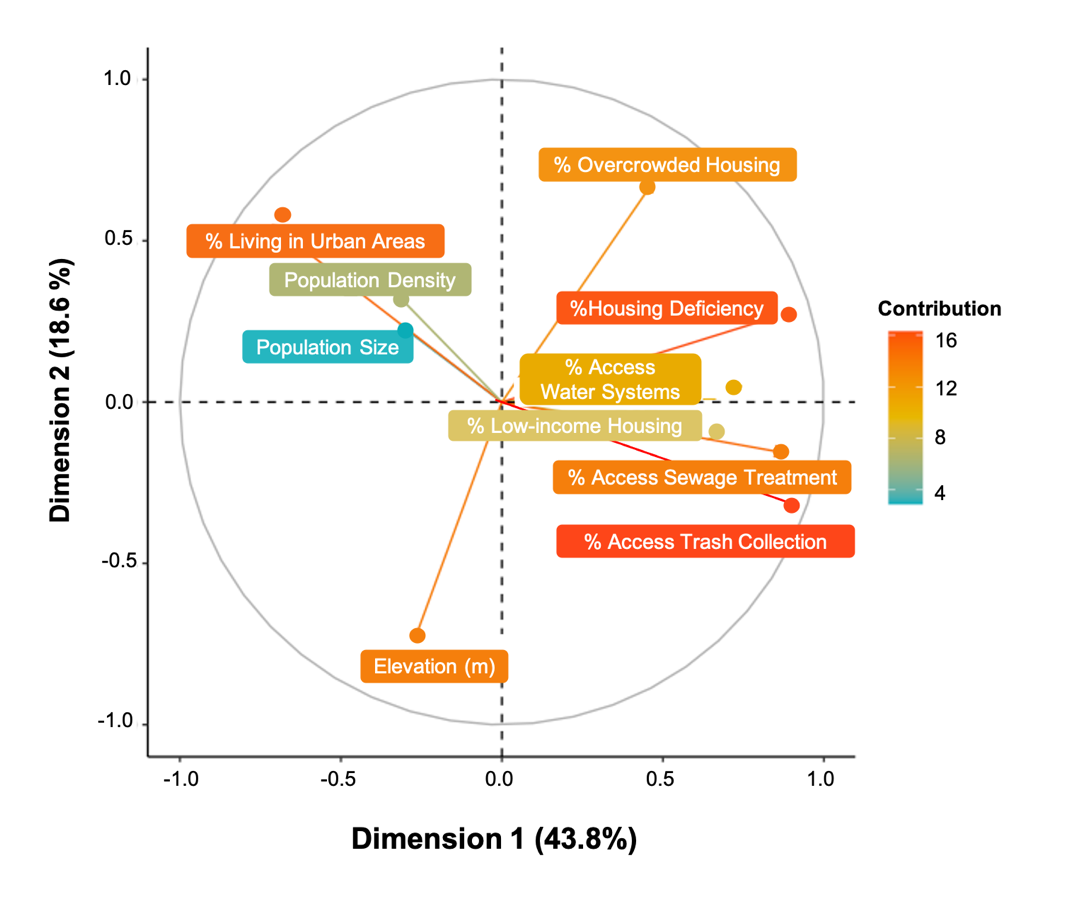

**Fig. S5. Principal components analysis of elevation and socio-economic features.**

The first component (contributing 43.8% to the explained variance) had a positive association with socio-economic indicators and a negative association with the percent of the municipality living in urban areas. The second component (contributing 18.6%) had a negative association with elevation and a positive association with percent of the municipality residing in overcrowded housing and the percent of the population living in urban areas. Finally, the third component (contributing 11.0%) had a strong negative association with the population size and population density.

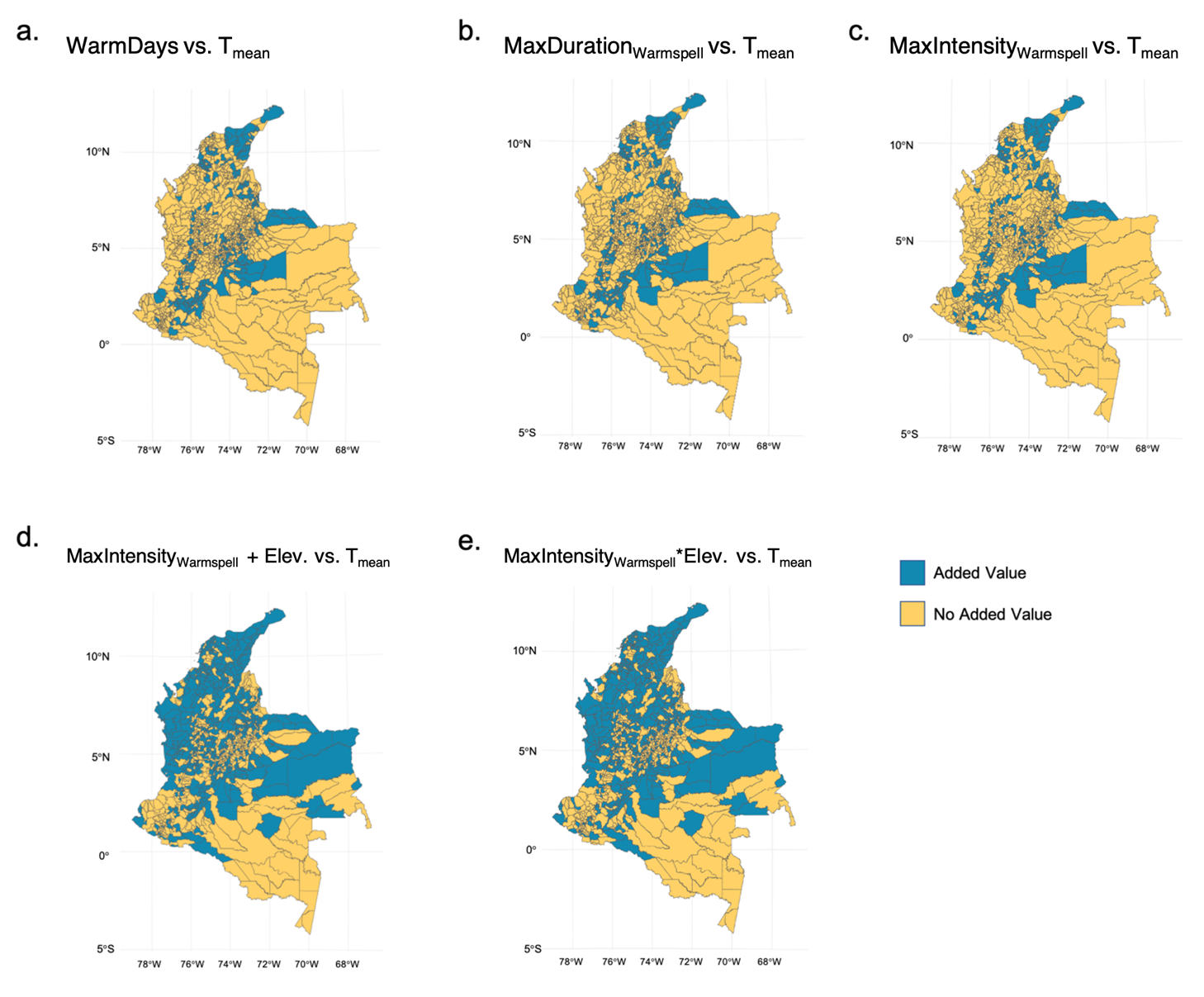

**Fig. S6. Added value of the extreme temperature models compared to mean temperature model.**

Difference in the mean absolute error (MAE) for the extreme temperature models compared to the MAE for the mean temperature model (including department-specific monthly random effects, year-specific spatial random effects, and regional fixed effect) for (a) the number of warm days, or days with temperatures exceeding the 90^th^ percentile of the monthly long-term average (T_90-mean_ threshold), (b) the maximum duration of a warmspell, (c) the maximum temperature intensity of a warmspell compared to the monthly long-term mean (MaxIntensity_Warmspell_), and the (d) additive and (e) interactive effects of MaxIntensity_Warmspell_ with the municipality’s elevation. Municipalities in blue have positive values, indicating that the extreme temperature model improves the model fit in these areas; whereas those in yellow did not have an improved model fit.

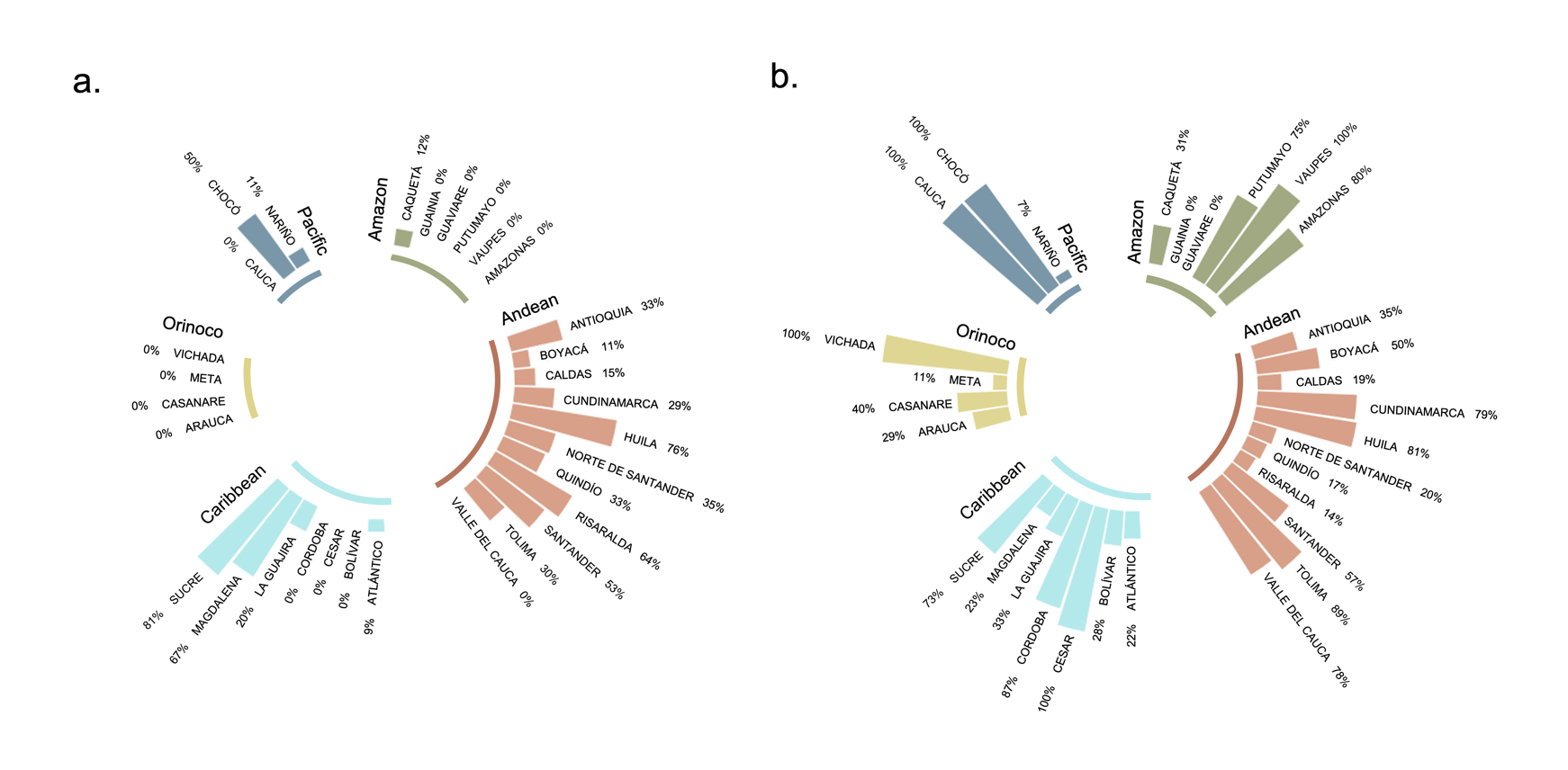

**Fig. S7. Proportion of municipalities per department with added value of MaxIntensity_Warmspell_ over T_mean_ model.**

**a.** Circular bar plots indicating the proportion of municipalities within each department that had a lower mean absolute error (MAE) for the MaxIntensity_Warmspell_ model compared to the T_mean_ model, indicating improved model predictions. Departments are grouped and colored according to the five bioclimatic regions, Amazon, Andean, Caribbean, Orinoco, and the Pacific. **b.** Proportion of municipalities within each department that had a lower mean absolute error (MAE) for the MaxIntensity_Warmspell_ *Elevation model compared to the T_mean_ model.

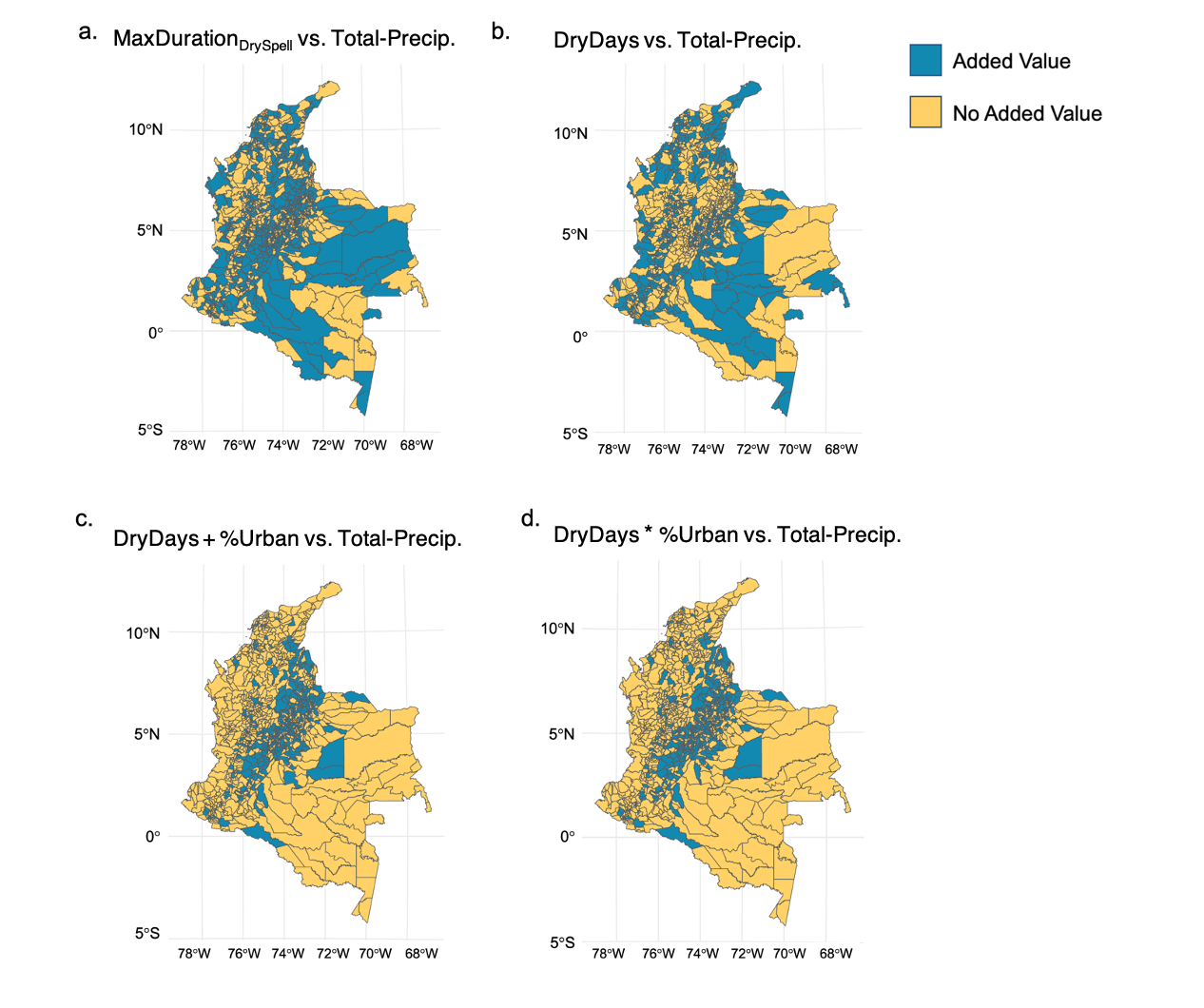

**Fig. S8. Added value of the extreme dry conditions compared to the total precipitation model.**

Difference in the mean absolute error (MAE) for the dry conditions models compared to the MAE for the total precipitation model (including department-specific monthly random effects, year-specific spatial random effects, and regional fixed effects) for (a) Maximum duration of days with < 1 mm precipitation (MaxDuration _DrySpell_), (b) the number of days with < 1 mm precipitation (DryDays), and the (c) additive and (d) interactive effects of DryDays with the percent of the municipality residing in urban areas (% Urban). Municipalities in blue have positive values, indicating that the dry condition model has an improved model fit in these areas; whereas municipalities in yellow do not have an improved model fit.

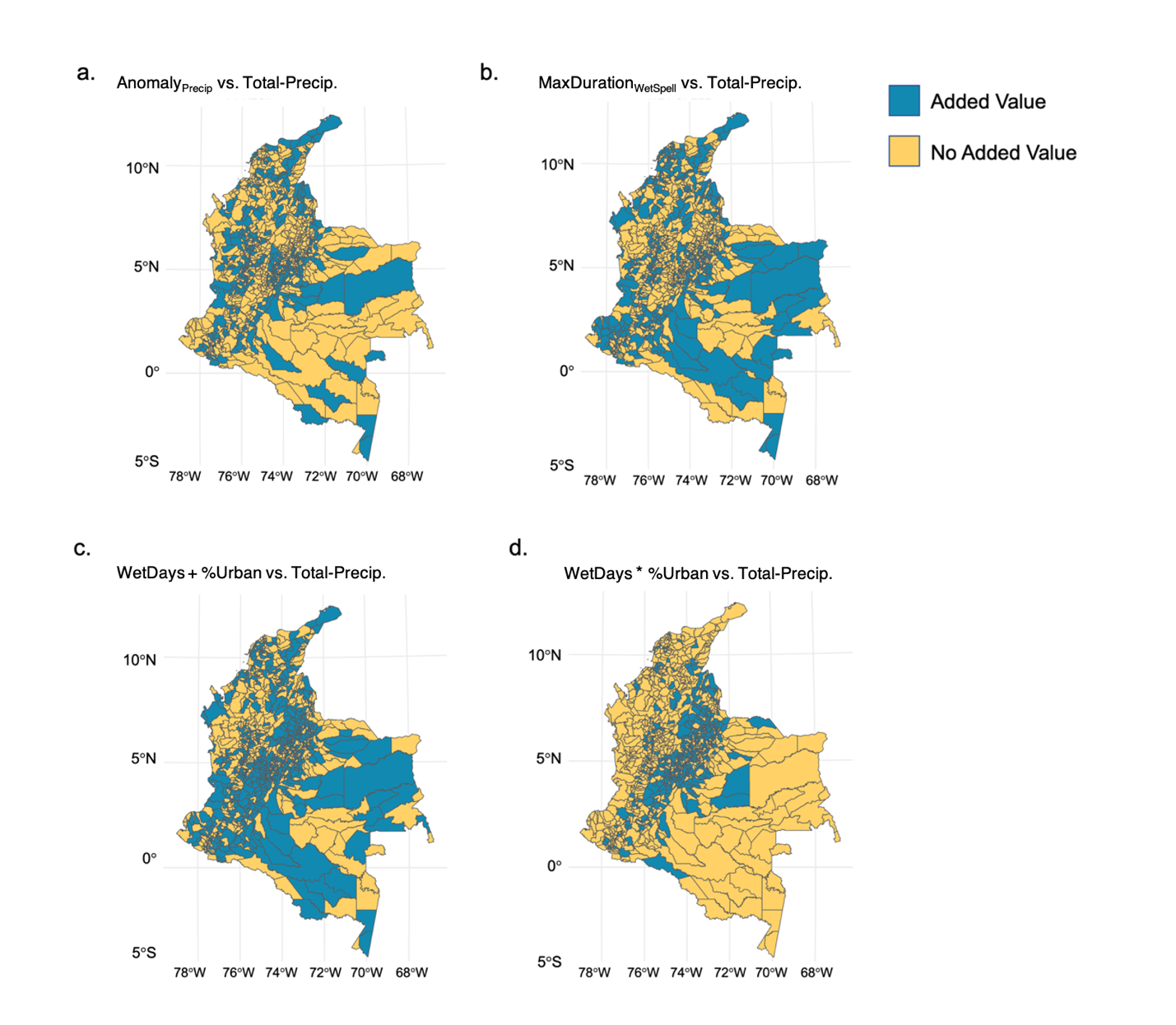

**Fig. S9. Added value of the extreme wet conditions compared to the total precipitation model.**

Maps visualizing the difference in mean absolute error (MAE) for the extreme wet conditions models compared to the MAE for the total precipitation model (including department-specific monthly random effects, municipality-specific yearly spatial random effects, and regional fixed effects). Municipalities in blue have positive values, indicating that the extreme wet conditions model has an improved model fit over total precipitation in these areas; whereas municipalities in yellow do not have an improved model fit. **a.** Comparison of the Anomaly_PRECIP_ model compared to the Total-Precip model. **b.** Comparison of the wet spell duration model vs. the Total-Precip model. **c.** Comparison of the WetDays model vs. the Total-Precip model. **d.** Comparison of the model with interactive effects of WetDays and the percent of the municipality residing in urban areas (% Urban) vs. the Total-Precip model.

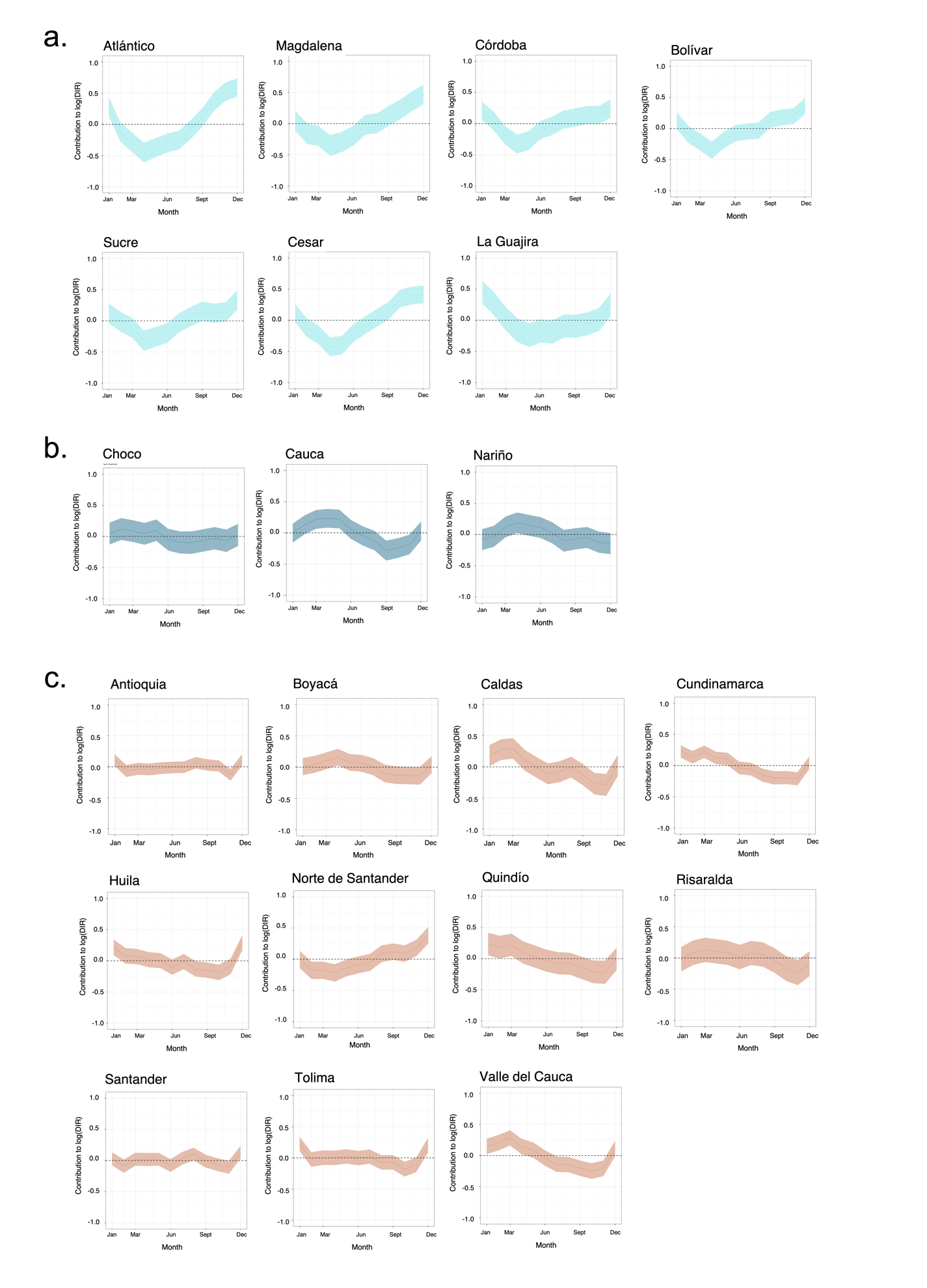

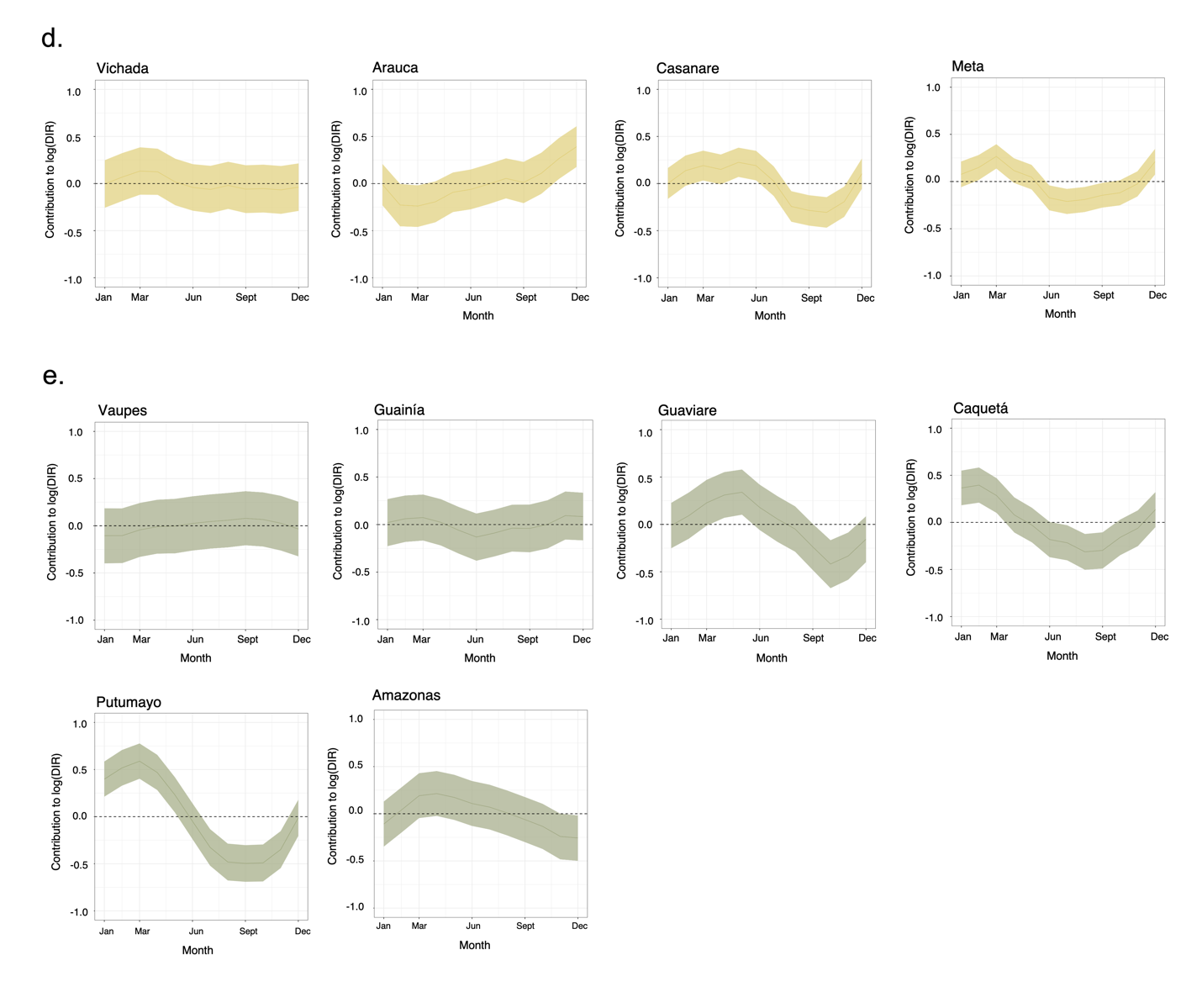

**Fig. S10. Posterior distributions of department-level monthly random effects**

Posterior mean (solid curve) and 95% credible interval (shaded area) of marginal posterior distribution of the department-level monthly random effects at the linear predictor scale from January to December for the 31 mainland departments in Colombia. This shows the contribution of the random effects to the log of the dengue incidence rate using the final extreme temperature*elevation—dry spell*urbanicity interaction model. Departments are grouped and colored according to their bioclimatic region **a.** Carribbean region. **b.** Pacific. **c.** Andes. **d.** Orinoco. **e.** Amazon

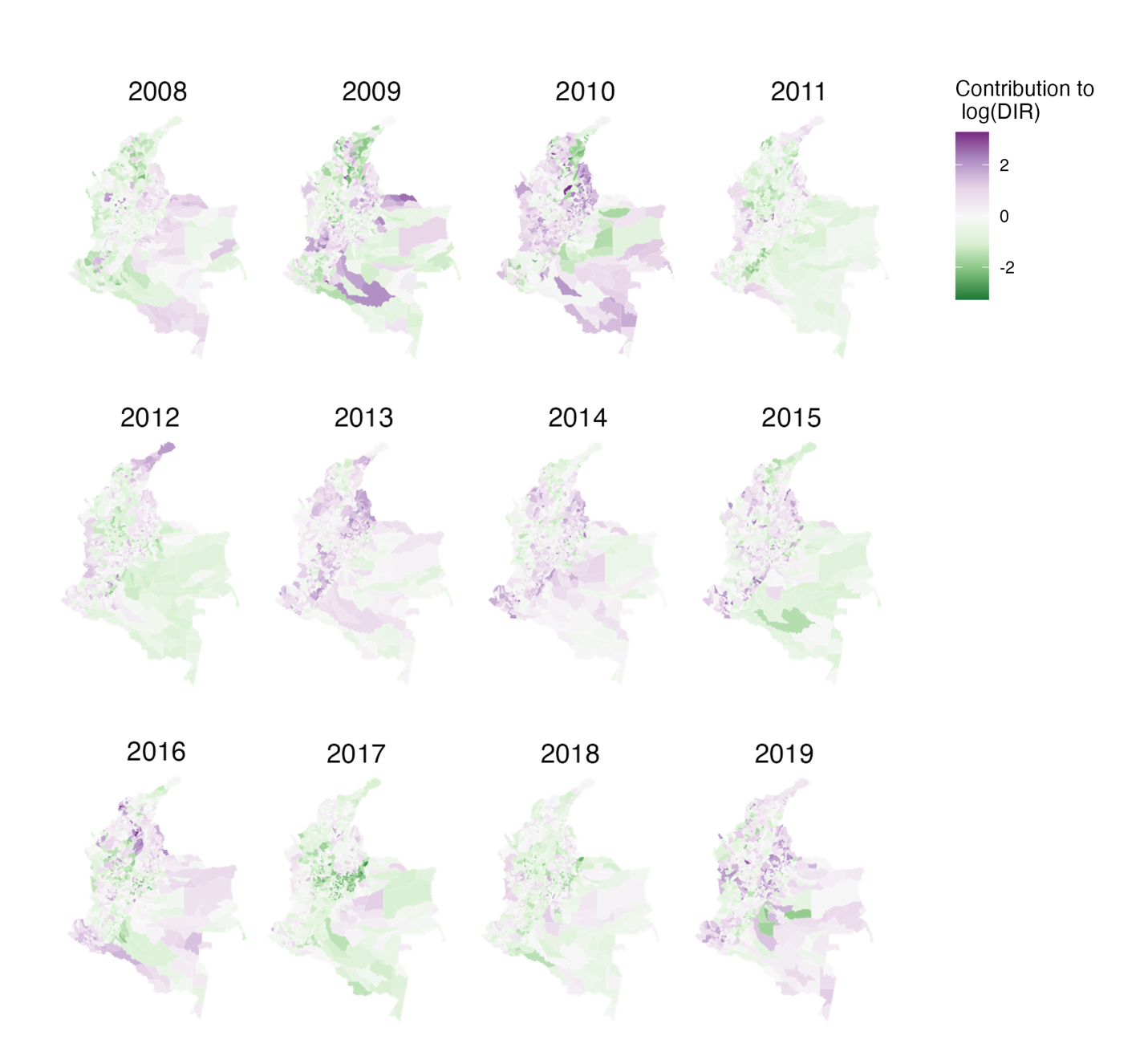

**Fig. S11. Contribution of yearly municipality-level spatial random effects to dengue incidence rate estimates**

Marginal posterior mean of the combined spatially structured and unstructured random effects at the linear predictor scale per year from 2008 to 2019. This shows the contribution of the municipality-level spatial random effects to the log of the dengue incidence rate for the final extreme temperature*elevation—dry spell*urbanicity interaction model.

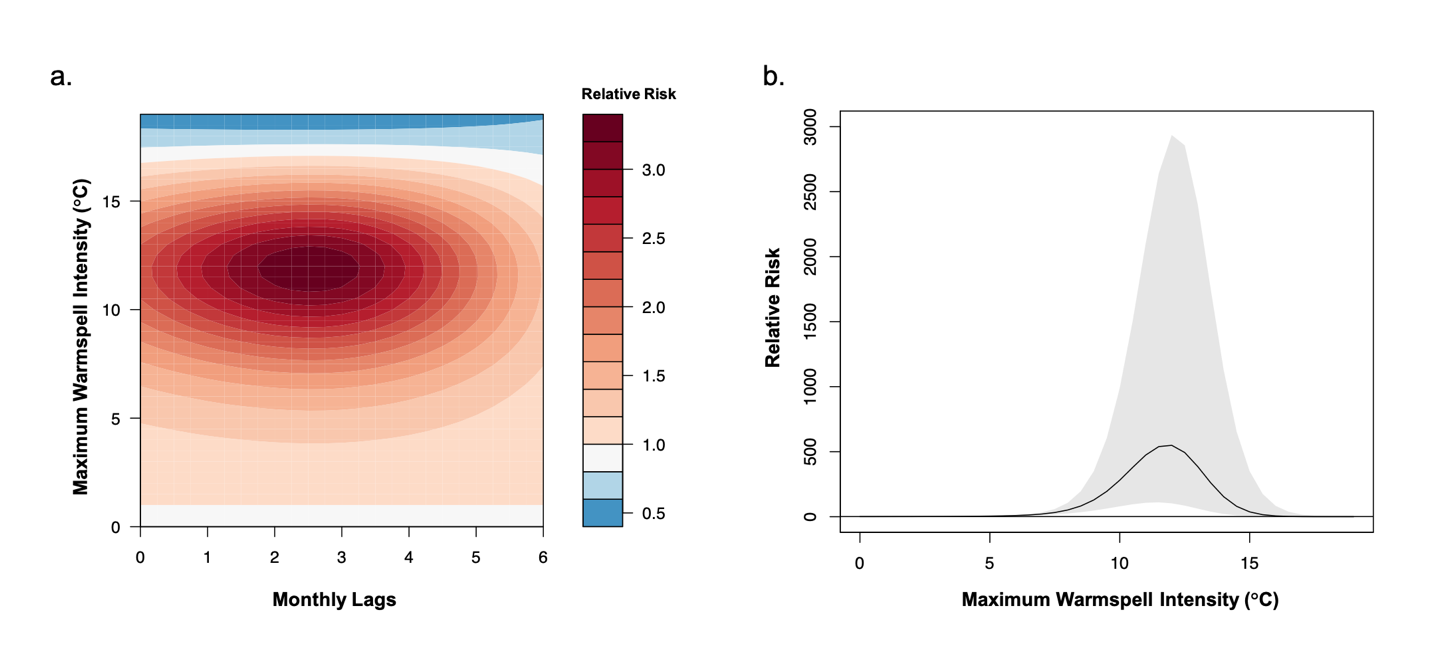

**Fig. S12. Effect of maximum warmspell intensity on dengue relative risk (RR) (included final model). a**. Contour plot of the exposure–lag–response association between monthly maximum warmspell intensity (MaxIntensity_Warmspell_)and dengue RR for 0 to 6-month lags, relative to the overall mean of the MaxIntensity_Warmspell_ for Colombia (1.1°C). The darker the red shading, the greater the increase in dengue RR compared to 1.1°C; the darker the blue shading, the greater the decrease in dengue RR compared to 1.1°C._._ **b**. Cumulative exposure–response associations for MaxIntensity_Warmspell_ and dengue RR relative to the overall mean of the MaxIntensity_Warmspell_ for Colombia (1.1°C), incorporating associations across all time lags (0 to 6-months). Shaded areas represent 95% Confidence Intervals. Note that MaxIntensity_Warmspell_ exceeded 10°C in only five instances during the study period (five municipalities in the Cauca Department in August 2012).

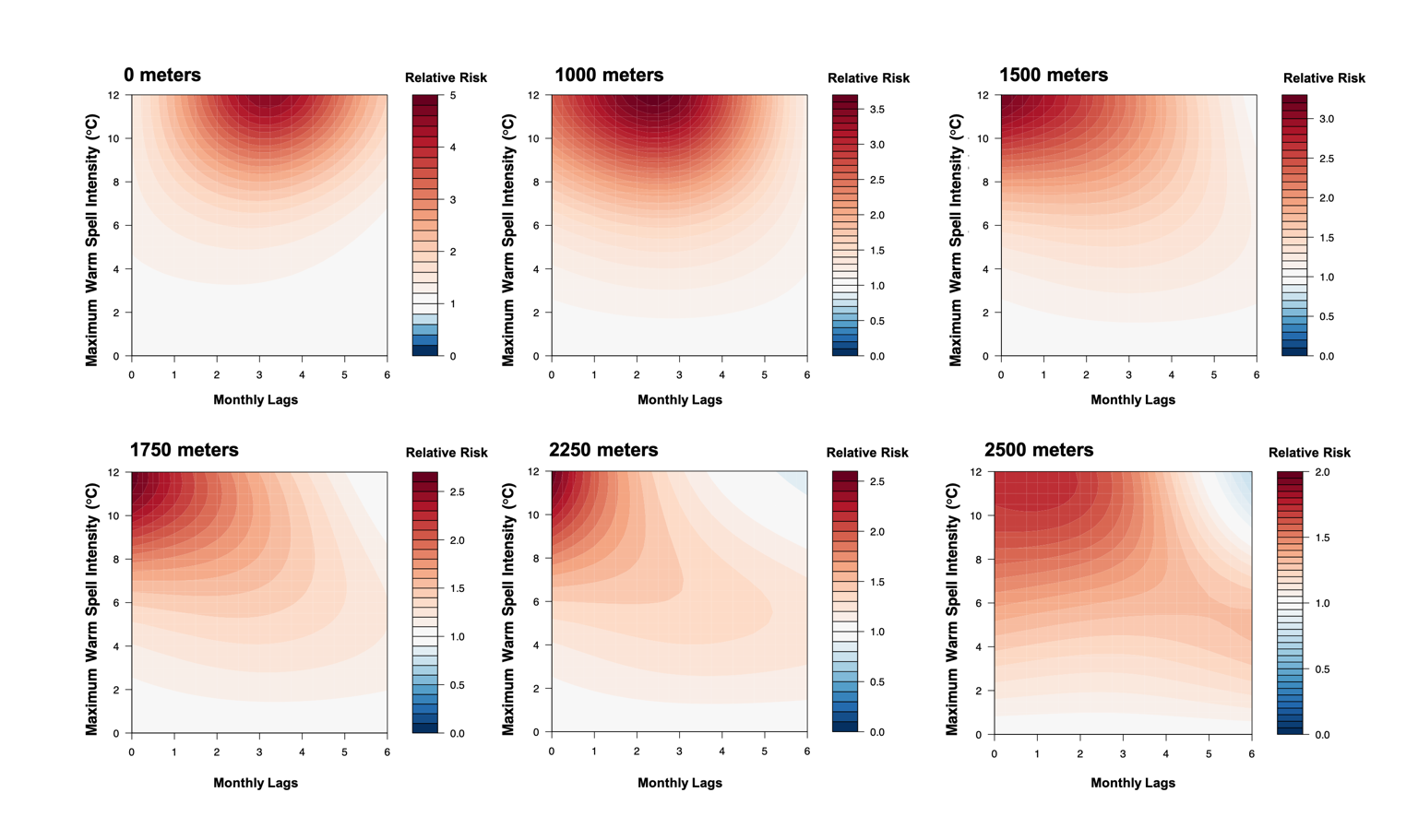

**Fig. S13. Association between maximum warmspell intensity and dengue risk at different elevations.** Contour plots of the exposure-lag-response association between the monthly maximum warmspell intensity (MaxIntensity_Warmspell_), compared to the overall mean of the MaxIntensity_Warmspell_ for Colombia (1.1°C) and dengue risk at different elevations. Red shading indicates a higher relative risk (RR) compared to mean temperature conditions; a RR < 1.0 indicates a decrease in risk.

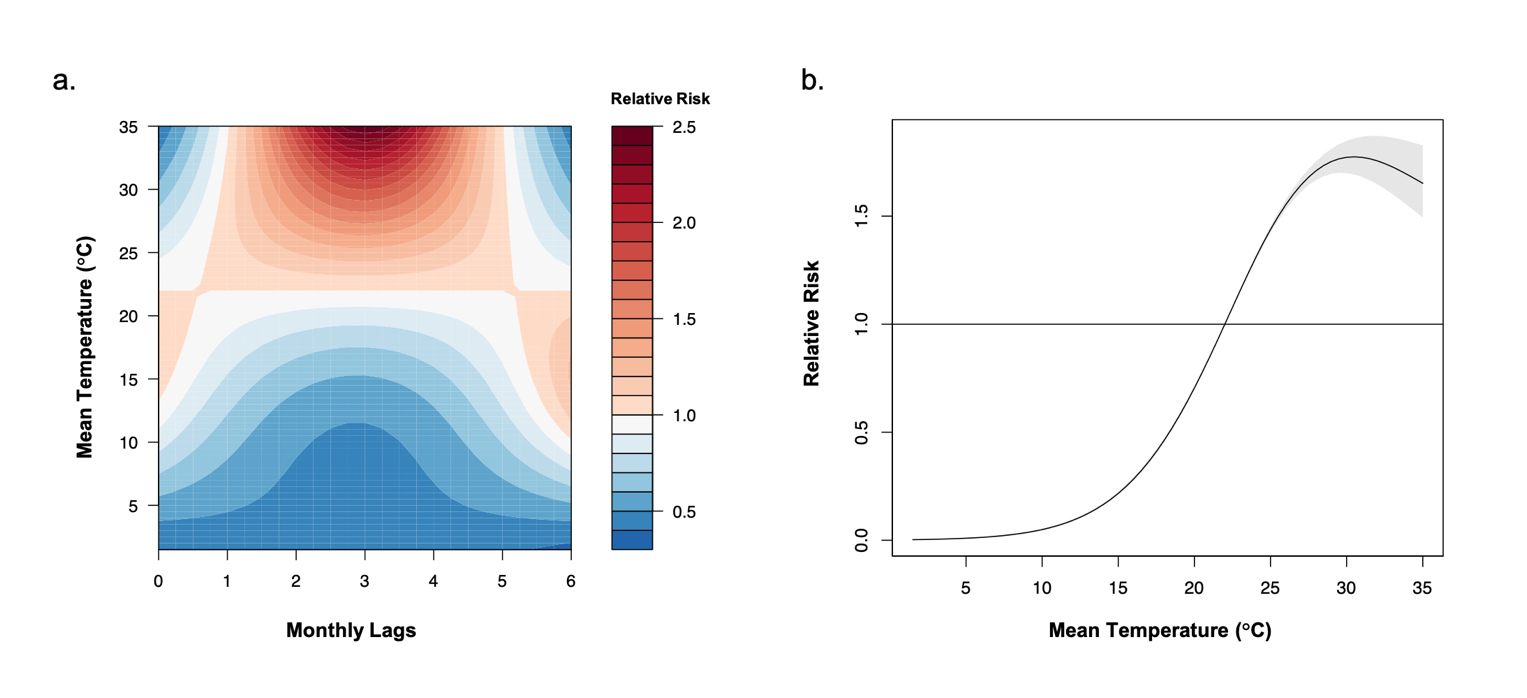

**Fig. S14. Effect of monthly T_mean_ on dengue relative risk (RR) (excluded from final model). a**. Contour plot of the exposure–lag–response association between T_mean_ and dengue RR for 0 to 6-month lags, relative to the overall mean T_mean_ for Colombia (22°C). The darker the red shading, the greater the increase in dengue RR compared to the overall mean T_mean_; the darker the blue shading, the greater the decrease in dengue RR compared to 22°C_._ **b**. Cumulative exposure–response associations for T_mean_ and dengue RR relative to the overall mean T_mean_ for Colombia (22°C), incorporating associations across all time lags (0 to 6-months). Shaded areas represent 95% Confidence Intervals (CI). The dengue RR peaked at a T_mean_ value of 30.5°C (95% CI: 29.5–32.0°C), with overlapping CIs for the range of mean optimal temperatures for *Ae. aegypti* DENV transmission reported via experimental studies (29.1°C; 95% CI: 28.4–29.8°C)^25^.

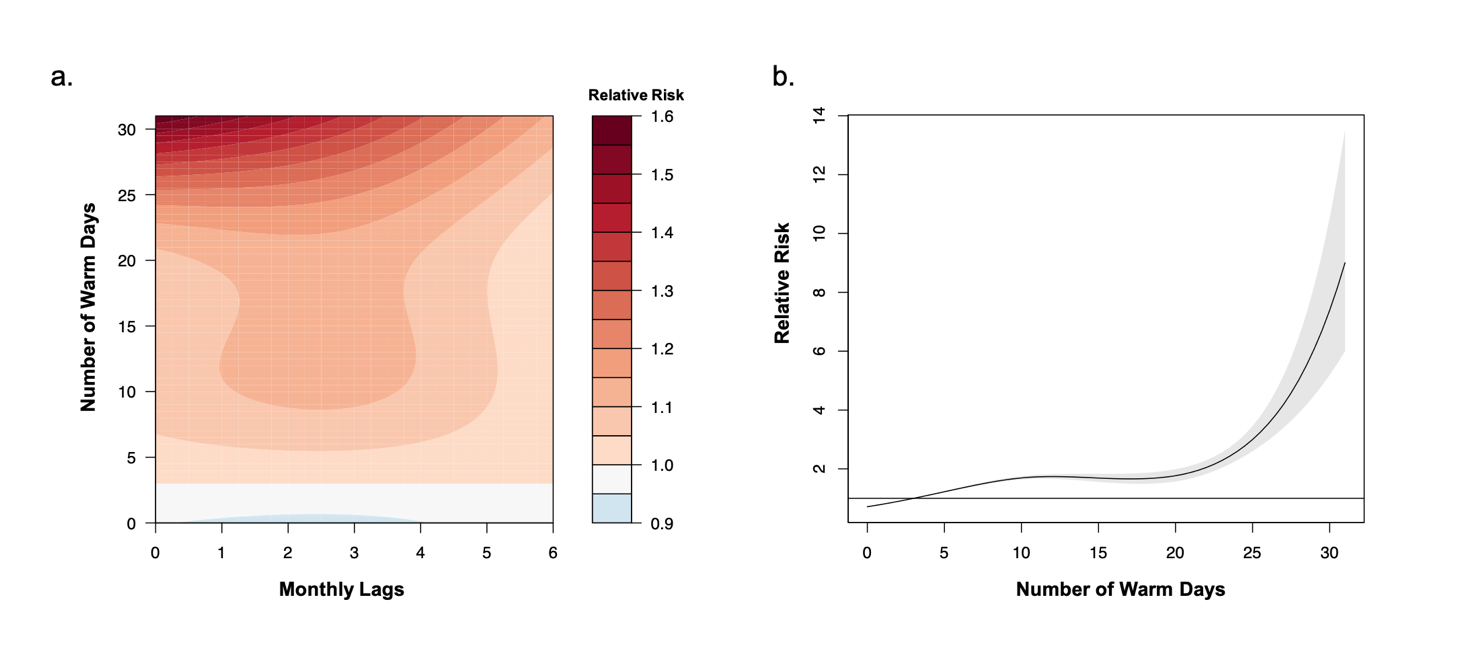

**Fig. S15. Effect of the number of warm days per month on dengue relative risk (RR) (excluded from final model). a**. Contour plot of the exposure–lag–response association for the number of warm days per month and dengue RR for 0 to 6-month lags, relative to the overall mean number of warm days per month for Colombia (2 days). The darker the red shading, the greater the increase in dengue RR compared to 2 warm days per month. **b**. Cumulative exposure–response associations for the number of warm days per month and dengue RR, relative to the overall mean number of warm days per month for Colombia (2 days), incorporating associations across all time lags (0 to 6-months). Shaded areas represent 95% Confidence Intervals (CI). The dengue RR exceeded 1.0 after 2 warm days per month and increases dramatically beyond 25 warms days per month (the corresponding 95% CI also widens beyond 25 warm days per month).

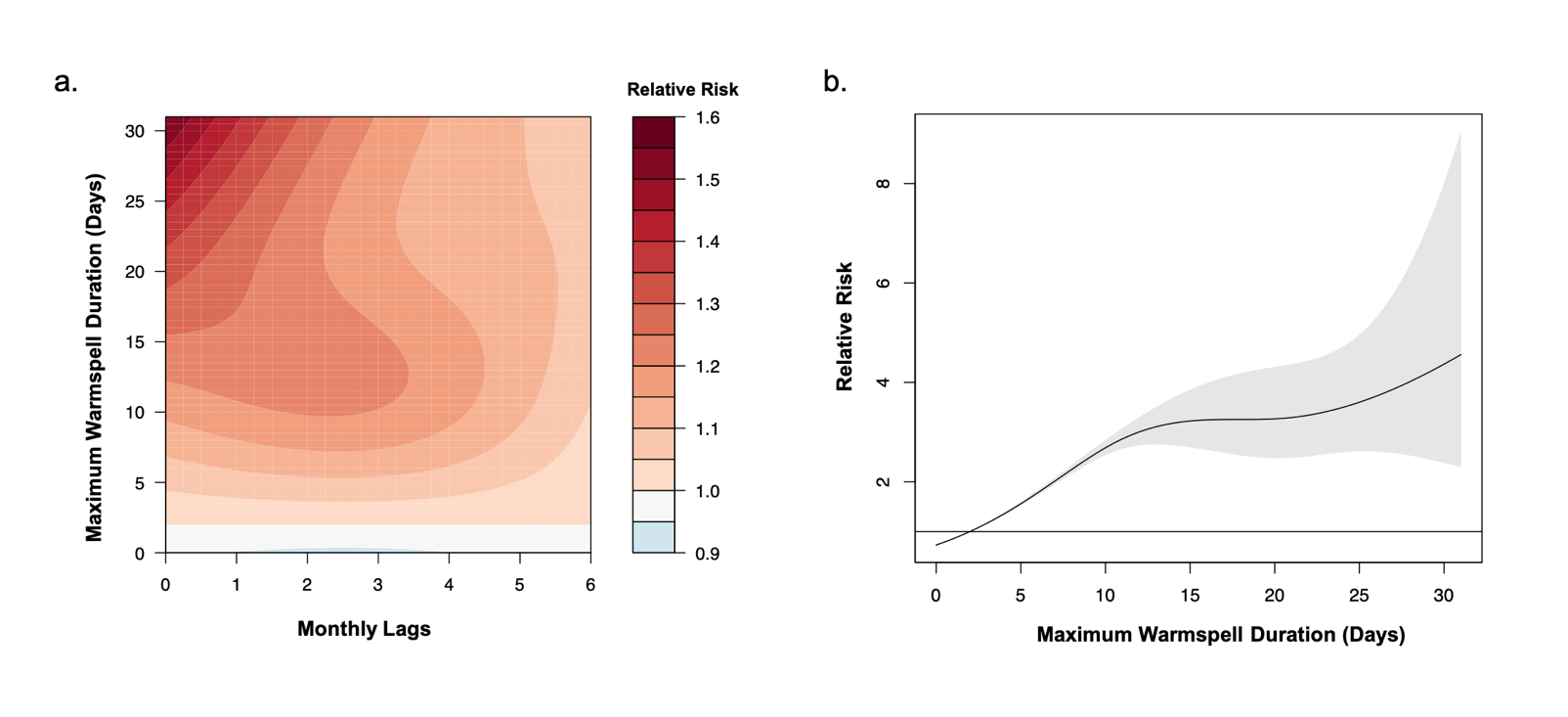

**Fig. S16. Effect of the monthly maximum warmspell duration on dengue relative risk (RR) (excluded from final model). a**. Contour plot of the exposure–lag–response association for maximum warmspell duration and dengue RR for 0 to 6-month lags, relative to the overall mean of the maximum warmspell duration per month for Colombia (2 days). The darker the red shading, the greater the increase in dengue RR compared to a warmspell of 2 days per month. **b**. Cumulative exposure–response associations for the maximum warmspell duration per month and dengue RR, relative to the overall mean of the maximum warmspell duration per month for Colombia (2 days), incorporating associations across all time lags (0 to 6-months). Shaded areas represent 95% Confidence Intervals (CI). The dengue RR exceeded 1.0 after a maximum warmspell duration of 2 days per month and increases non-linearly for a 10-day or longer maximum warmspells. The corresponding 95% CI also widens beyond a 10-day maximum warmspell, with 2.1% of the total dataset (N=3459/161,280 records) having maximum warmspell durations of more than 10 days.
